# Uncovering Phenotypic Expansion in AXIN2-Related Disorders through Precision Animal Modeling

**DOI:** 10.1101/2024.12.05.24318524

**Authors:** Nathalie M. Aceves-Ewing, Denise G. Lanza, Paul C. Marcogliese, Di Lu, Chih-Wei Hsu, Matthew Gonzalez, Audrey E. Christiansen, Tara L. Rasmussen, Alex J. Ho, Angelina Gaspero, John Seavitt, Mary E. Dickinson, Bo Yuan, Brian J. Shayota, Stephanie Pachter, Xiaolin Hu, Debra Lynn Day-Salvatore, Laura Mackay, Oguz Kanca, Michael F. Wangler, Lorraine Potocki, Jill A. Rosenfeld, Richard Alan Lewis, Hsiao-Tuan Chao, Brendan Lee, Sukyeong Lee, Undiagnosed Diseases Network, Baylor College of Medicine Center for Precision Medicine Models, Shinya Yamamoto, Hugo J. Bellen, Lindsay C. Burrage, Jason D. Heaney

**Affiliations:** Department of Molecular and Human Genetics, Baylor College of Medicine (BCM), Houston, TX, 77030, USA; Department of Biochemistry and Medical Genetics, Rady Faculty of Health Sciences, University of Manitoba, Winnipeg, MB, R3E 0J9, Canada; Children’s Hospital Research Institute of Manitoba (CHRIM), Winnipeg, MB, R3E 3P4, Canada; Jan and Dan Duncan Neurological Research Institute, Texas Children’s Hospital (TCH), Houston, TX, 77030, USA; Department of Integrative Physiology, Baylor College of Medicine, Houston, TX, 77030, USA; Optical Imaging and Vital Microscopy Core, Baylor College of Medicine, Houston, TX, 77030, USA; Verna and Marrs McLean Department of Biochemistry and Molecular Pharmacology, Baylor College of Medicine, Houston, TX, 77030, USA; Advanced Technology Core for Macromolecular X-ray Crystallography, Baylor College of Medicine, Houston, TX, 77030, USA; Human Genome Sequencing Center, Baylor College of Medicine, Houston, TX, 77030, USA; Division of Medical Genetics, Department of Pediatrics, University of Utah, Salt Lake City, UT, 84108, USA; Atlantic Health System, Summit, NJ 07901, USA; GeneDX, Gaithersburg MD, 20877, USA; Department of Medical Genetics and Genomic Medicine, Saint Peter’s University Hospital, New Brunswick, NJ, 08901, USA; Department of Pediatrics, Division of Genetics and Metabolism, University of Texas Southwestern Medical Center, Dallas, TX, USA; Texas Children’s Hospital, Houston, TX, 77030, USA; Department of Neuroscience, BCM, Houston, TX, 77030, USA; The Cain Pediatric Neurology Research Foundation Laboratories, Jan and Dan Duncan Neurological Research Institute, Texas Children’s Hospital, Houston, Texas, USA; McNair Medical Institute, The Robert and Janice McNair Foundation, Houston, Texas, USA; Division of Neurology and Developmental Neuroscience, Department of Pediatrics, Baylor College of Medicine, Houston, Texas, USA; Member list in supplemental material; Dan L. Duncan Comprehensive Cancer Center, Baylor College of Medicine, Houston, TX, 77030, USA

**Author notes:** These authors contributed equally (Nathalie M. Aceves-Ewing, Denise G. Lanza, Paul C. Marcogliese, Hugo J. Bellen, Lindsay C. Burrage, Jason D. Heaney).

**Keywords:** ectodermal dysplasia, WNT/β-catenin signaling, oligodontia-colorectal cancer, AXIN2, tankyrase-binding domain

## Abstract

Heterozygous pathogenic variants in *AXIN2* are associated with oligodontia-colorectal cancer syndrome (ODCRCS), a disorder characterized by oligodontia, colorectal cancer, and in some cases, sparse hair and eyebrows. We have identified four individuals with one of two *de novo*, heterozygous variants (NM_004655.4:c.196G>A, p.(Glu66Lys) and c.199G>A, p.(Gly67Arg)) in *AXIN2* whose presentations expand the phenotype of AXIN2-related disorders. In addition to ODCRCS features, these individuals have global developmental delay, microcephaly, and limb, ophthalmologic, and renal abnormalities. Structural modeling of these variants suggests that they disrupt AXIN2 binding to tankyrase, which regulates AXIN2 levels through PARsylation and subsequent proteasomal degradation. To test whether these variants produce a phenotype *in vivo*, we utilized an innovative prime editing N1 screen to phenotype heterozygous (p.E66K) mouse embryos, which were perinatal lethal with short palate and skeletal abnormalities, contrary to published viable *Axin2* null mouse models. Modeling of the p.E66K variant in the *Drosophila* wing revealed gain-of-function activity compared to reference AXIN2. However, the variant showed loss-of-function activity in the fly eye compared to reference AXIN2, suggesting that the mechanism by which p.E66K affects AXIN2 function is cell context-dependent. Together, our studies in humans, mice, and flies demonstrate that specific variants in the tankyrase-binding domain of AXIN2 are pathogenic, leading to phenotypic expansion with context-dependent effects on AXIN2 function and WNT signaling. Moreover, the modeling strategies used to demonstrate variant pathogenicity may be beneficial for the resolution of other *de novo* heterozygous variants of uncertain significance associated with congenital anomalies in humans.

## Introduction

The canonical WNT signaling pathway, also referred to as the β-catenin-dependent WNT pathway, is evolutionarily conserved and critical to embryonic development and adult homeostasis in nearly all animals. Since the WNT/β-catenin pathway is involved in many early developmental processes and in the maintenance of cell proliferation, diseases associated with dysregulated WNT signaling include a wide range of Mendelian and non-Mendelian phenotypes and several types of cancer. Some of the known disorders associated with aberrant WNT signaling include tooth agenesis^1-4^, neurodegenerative disorders^5-7^, and renal disease^8^. Cancers associated with members of the WNT signaling pathway include breast cancer^9-11^, hepatocellular carcinoma^11-13^, and colorectal cancer^1-3,11,13-17^.

The inhibition and activation of the canonical WNT/β-catenin pathway involves multiple extracellular, membrane-bound, cytoplasmic, and nuclear proteins^18-22^. In the absence of WNT ligand binding to the receptor complex consisting of FZD and LRP5/6, β-catenin is constitutively phosphorylated by the destruction complex (composed of APC, CK1, GSK3β, and AXIN1 or AXIN2), leading to its ubiquitination and proteasomal degradation^21,23-28^. However, upon WNT binding, the activity of the destruction complex is inhibited and accumulated β-catenin translocates to the nucleus, where it associates with TCF/LEF transcription factors to activate the expression of genes involved in cell fate determination, cell proliferation, and other key cellular processes^14,26,27^. Critically, in some cell contexts AXIN2 can also positively regulate WNT signaling by binding to and maintaining the phosphorylation of the LRP5/6 receptors after WNT activation of its receptor complex^29-32^.

Somatic variants in *AXIN2* have been associated with somatic colorectal cancer (MIM #114500) whereas germline variants in *AXIN2* have been associated with oligodontia-colorectal cancer syndrome (ODCRCS, MIM #608615)^1-4,9,10,15,16,33^. ODCRCS most commonly includes phenotypes such as oligodontia, colon polyps, colorectal cancer, and sparse hair and eyebrows^1-4,9,10,15,16,33,34^. To date, most published pathogenic variants in *AXIN2* cluster towards the C-terminus of the protein as well as a few variants in the Regulator of G protein signaling (RGS), GSK3β-binding, and β-catenin-binding domains^1-4,9,15,16^. However, to date, no pathogenic variants have been identified in the tankyrase (TNKS)-binding domain^1-4,15,16,35-37^. This domain is necessary for TNKS binding, which leads to the poly-ADP-ribosylatation (PARsylation), ubiquitination, and proteasomal degradation of AXIN2^35,37^.

In this study, we report three individuals with ectodermal dysplasia, musculoskeletal anomalies, genitourinary abnormalities, and a *de novo* heterozygous variant in the TNKS-binding domain (p.Glu66Lys, referred to as E66K) of AXIN2. As pathogenic variants in this domain have not been previously reported and as the phenotype associated with these variants was hypothesized to be lethal in mice, we combined structural and fly modeling with innovative mouse modeling strategies to demonstrate the pathogenicity of the p.E66K variant. In addition, we used amino acid conservation and population databases to identify a neighboring variant (p.Gly67Arg, referred to as G67R) that causes an overlapping human phenotype. Overall, we demonstrate that variants impacting key residues in the TNKS-binding domain of AXIN2 represent a phenotypic expansion and novel ectodermal dysplasia associated with congenital anomalies.

## Materials and Methods

### Clinical Study

Participant 1 and both parents were enrolled in the Baylor College of Medicine (BCM) site for the Undiagnosed Diseases Network (UDN). This study was approved by the Institutional Review Board (IRB) at both the National Institutes of Health and BCM. The family provided written informed consent prior to the initiation of any research procedures. Additional cases (participants 2, 3, and 4) were identified through a search of ClinVar^38^ for variants at the same or nearby positions and communications with the submitters of these variants (GeneDx and Invitae). Participants 2, 3, and 4 and their families also provided written informed consent prior to the initiation of research procedures under an IRB-approved protocol at BCM. *As per medRxiv policy, the case histories for the participants and their images have been removed. To obtain more detailed information, please contact the authors*.

### Exome and Genome Sequencing

For participant 1, clinical trio exome sequencing was performed at Baylor Genetics as previously described^39^ and transferred to the BCM UDN site for research re-analysis. Trio genome sequencing was performed through the UDN Sequencing Core at Baylor Genetics as previously described^40^, and research reanalysis was performed at the BCM UDN site. The research exome and genome re-analyses prioritized rare, *de novo* and biallelic variants that had an allele frequency of < 1% in gnomAD v4.0.0^41^ and in the BCM UDN internal sequencing database (> 1200 samples). Codified Genomics was used for variant filtering and prioritization. Participant 2’s variant was detected on an Invitae Hereditary Colorectal Cancer panel as part of an evaluation for colon polyps. This next generation sequencing panel included 20 genes associated with predisposition to colorectal cancer. Sanger sequencing was performed using DNA extracted from parental buccal samples at Baylor Genetics Laboratory to confirm that the *AXIN2* variant identified in participant 2 is *de novo*. Participants 3 and 4 had clinical trio exome sequencing performed through GeneDx. For CADD Scores, CADD GRCh37-v1.7 was used.

### Conservation alignments

The conservation alignments were made using default parameters in Clustal Omega (CLUSTAL O(1.2.4) multiple sequence alignment)^42-44^ with *Homo sapiens* AXIN2 (Q9Y2T1) as the reference sequence aligned with *Mus musculus* AXIN2 (O88566), and *Homo sapiens* (O15169) and *Mus musculus* (O35625) AXIN1. Amino acid sequences were acquired from UniProt^45^. A similar alignment was performed using *Homo sapiens* AXIN2 (Q9Y2T1) as the reference sequence aligned with fly (*Drosophila melanogaster*) AXN (Q9V407) and AXIN2 amino acid sequences from the following organisms: *Mus musculus* (O88566), *Rattus norvegicus* (O70240), *Pan troglodytes* (K7CHS6), *Gorilla gorilla gorilla* (G3QN00), *Macaca mulatta* (A0A5F8AE66), *Pongo abelii* (H2NUH3), *Papio anubis* (A0A096NTX5), *Chlorocebus sabaeus* (A0A0D9QVR2), *Otolemur garnettii* (H0WYQ7), *Microcebus murinus* (A0A8B7F524), *Ictidomys tridecemlineatus* (I3N719), *Canis lupus familiaris* (A0A8C0YRK8), *Felis catus* (M3XAL0), *Mustela putorius furo* (M3YT87), *Ailuropoda melanoleuca* (G1LZK8), *Equus caballus* (F6SK96), *Pteropus vampyrus* (A0A6P6CVR3), *Bos taurus* (G3X7P3), *Sus scrofa* (A0A287BFQ6), *Tursiops truncates* (A0A6J3QL77), *Sarcophilus harrisii* (G3VT89), *Monodelphis domestica* (F6TVK6), *Anolis carolinensis* (H9G5B4), *Pelodiscus sinensis* (K7G9P5), *Gallus gallus* (A0A1D5PPH8), *Latimeria chalumnae* (H3BCV4), *Ficedula albicollis* (U3JIC1), *Danio rerio* (P57095), *Oreochromis niloticus* (I3J624), *Xiphophorus maculatus* (A0A3B5QRG9), *Oryzias latipes* (H2L981), *Tetraodon nigroviridis* (H3C937), *Gasterosteus aculeatus* (G3Q3K5), *Xenopus tropicalis* (A0A803JRP4), *Astyanax mexicanus* (A0A3B1KDC5), and *Takifugu rubripes* (H2SP31).

### Structural modeling

The side chains of the mutated E66K and G67R residues in the TNKS binding motif 2 (TBM2) of AXIN2 were modeled based on the crystal structure of the TNKS ankyrin-repeat clusters 2-3 (ARC2-3) and AXIN1 complex (PDB ID: 3UTM) as a template^37^. For each variant, a rotamer conformation was selected for the mutated residue to model the side chain, followed by minor adjustments of the *chi* angles. Structural modelling was performed using Coot^46^.

### Mouse colony management

All mice were housed in microisolator cages in ventilated racks within the ABBR animal facility at BCM. The rooms were temperature controlled and humidity controlled with a 6:00 AM to 8:00 PM light:dark cycle. The mice were given *ad libitum* access to food (Lab Diet 5V5R) and autoclaved drinking water for the duration of the study. All studies were approved by the BCM Institutional Animal Care and Use Committee.

### *Axin2* p.E66K founder embryo screen

To generate founder embryos harboring a knock-in (KI) of the p.E66K (GAG>AAG) point mutation, as well as a p.G67 (GGG>GGC) synonymous mutation, two different design strategies were implemented, each using a single guide RNA (sgRNA) with a complementary single-stranded oligodeoxynucleotide (ssODN) repair donor for CRISPR/Cas9-initiated homology-directed repair (**Figure S1**). Inclusion of the synonymous mutation in the ssODN sequence alters the sgRNA target sequence, thus reducing guide affinity and suppressing secondary CRISPR genome editing events after homology-directed repair. The sgRNAs were selected by the BCM Genetically Engineered Rodent Model (BCM GERM) Core using the Wellcome Sanger Institute Genome Editing (WGE) website^47^. Complementary to the strand of the target site, corresponding ssODNs were designed with asymmetric homology arms flanking the p.E66K mutation relative to the predicted cut site of the gRNA. Sequences and genomic coordinates can be found in **Table S1**. The sgRNAs were synthesized by Synthego and custom ssODNs (Ultramers) were produced by Integrated DNA Technologies (Coralville, IA). The BCM GERM Core electroporated a mixture consisting of precomplexed sgRNA (6.6 µM) and Cas9 Protein (6 µM, PNA Bio), ssDNA mRNA (100 pmol) in a final volume of 10 μL 1xTE (RNAse-free) into at least 60 pronuclear stage C57BL/6J (Jackson Lab Strain #:000664; RRID:IMSR_JAX:000664) zygotes as previously described^48^. Embryos were transferred to pseudopregnant females the same day as electroporation and allowed to gestate for 18 days prior to sacrifice of the recipient dam and collection of embryos for subsequent genotyping and imaging.

### *Axin2^E66K/+^* constitutive knock-in mouse model

To generate a constitutive knock-in (KI) mouse model with the p.E66K (GAG>AAG) point mutation as well as a p.G67 (GGA>GGG) synonymous mutation, RNA-guided nuclease prime editing was utilized (**Figure S2A)**. A prime editing guide RNA (pegRNA) and a complementary sgRNA to facilitate DNA nicking were selected by the BCM GERM Core using Peg Finder^49^. Again, the synonymous mutation was included to suppress secondary genome editing events. Target sequences, genomic coordinates, and guide sequences can be found in **Table S2 and S3**. The pegRNA was purchased as a custom gRNA from Integrated DNA Technologies (Coralville, IA) and the nicking gRNAs as synthesized sgRNAs from Synthego (Redwood City, CA). Prime editor (PE2) mRNA was generated from linearized plasmid DNA (pCMV-PE2, addgene.org/132775/) using the mMESSAGE mMACHINE T7 ULTRA Transcription Kit (ThermoFisher, AM1345). The BCM GERM Core microinjected a mixture consisting of pegRNA (75ng/μL), sgRNA (75ng/μL), and PE2 mRNA (150ng/μL) in a final volume of 50 μL 1xTE (RNAse-free) into the cytoplasm of C57BL/6J (Jackson Lab Strain #:000664; RRID:IMSR_JAX:000664) pronuclear stage zygotes as previously described^50^. DNA was isolated from ear punches and PCR was performed with AmpliTaq Gold 360 Master Mix (Applied Biosystems, 4398881) using primers Axin2 E2 F (5’ TCCAGAGAGGAGGCTCACAT) and Axin2 E2 R (5’ AAACATGACCGAGCCGATCT). PCR products were Sanger sequenced and Synthego ICE analysis identified a founder male with approximately 20% mosaicism for an allele harboring the p.E66K point mutation and p.G67 synonymous mutation, as well as a p.S70 (TCG>TCC) synonymous mutation (**Figure S2B)**. This founder was used for breeding and subsequently sperm cryopreservation using the CARD method^51,52^. The allele is registered at Mouse Genome Informatics (MGI) as *Axin2^em1Bay^*(MGI:7768058).

### Embryo phenotyping

For the p.E66K embryo founder screen, embryos were collected at embryonic day (E)18.5, fixed in 4% paraformaldehyde (PFA), and stored at 4°C until they could be processed for μCT imaging, as previously described^53^. To generate N1 embryos harboring a *Axin2^E66K/+^* constitutive knock-in allele, timed matings between the *Axin2^E66K/+^* mosaic male founder and wild-type C57BL/6J females or *in vitro* fertilization with mosaic male sperm and wild-type C57BL/6J oocytes were used as previously described^52,54,55^. Timed matings were set up between male and female mice during the afternoon. If a vaginal plug was confirmed the next morning, female and male mice were separated, and embryos were stated to be at E0.5. Embryos were analyzed upon dissection at E18.5 with light microscopy. For μCT imaging, E18.5 embryos were fixed and processed as described for the founder screen. Embryos were scanned with a Bruker SkyScan 1272 μCT Scanner, and images were analyzed with Slicer (v5.2.2). Annotation of phenotypes was performed blinded to genotypes.

### Embryo genotyping

DNA was extracted from yolk sacs for PCR-based genotyping and Sanger sequencing. PCR was performed with AmpliTaq Gold 360 Master Mix using primers Axin2 E2 F (5’ TCCAGAGAGGAGGCTCACAT) and Axin2 E2 R (5’ AAACATGACCGAGCCGATCT). For the founder embryo screen, PCR products were Sanger sequenced, and Synthego Inference of CRISPR Edits (ICE) analysis was used to determine wild-type, insertion/deletion (non-homologous end joining), and variant (homology-directed repair) allele contribution in each embryo. For N1 embryos derived from the mosaic *Axin2^E66K/+^* constitutive knock-in founder male, the same primers were used to generate PCR products, which were subsequently digested with StyI High-Fidelity enzyme (NEB, R3500S). The expected size for the wild-type product is 653 bp; StyI cleaves at the p.E66K sequence in the *Axin2^E66K^* allele, resulting in 335 bp and 318 bp products (**Figure S2A&C**). Transmission of the *Axin2^E66K/+^*constitutive knock-in allele from the mosaic founder to N1 embryos was also confirmed via Sanger sequencing (**Figure S2B**).

### Generation of AXIN2 transgenic flies

*UAS-AXIN2* reference and variant transgenic flies were generated as previously described^56^. Specifically, the Gateway compatible *AXIN2* (NM_004655.4) open-reading frame (ORF) in the DONR221 vector was shuttled to the pGW-HA.attb^57^ UAS destination vector by LR clonase II (Invitrogen) by manufacturer protocol. The ORF harbors an endogenous stop codon, thus this closed clone will not be tagged with HA. Variants were generated via site-directed mutagenesis with the Q5 mutagenesis kit (NEB) followed by verification of the ORF by Sanger sequencing. Primers used for site-directed mutagenesis for the *AXIN2* p.E66K, R656X, and W663X variants are listed in **Table S4**. Final UAS constructs were microinjected into embryos containing the VK37 (PBac{y[+]-attP}VK00037) genomic docking site and integrated by ϕC31 mediated transgenesis^58^.

### Generation of *AxnTG4* mutant flies

The *Axn^TG4^* (*Axn^CR70218-TG4.2^*; BL_ 93803 – BDSC) allele was generated in a previous study as part of the large-scale gene disruption project^59^. Briefly, we generated homology donor intermediates to target an intron separating two exons in the coding region of *Axn*. Construct with homology arms (200 bases upstream and downstream of the target site) and *Axn*-specific sgRNA that targets the site (TCATCGAGTCAGTGGTGTAGAGG) was commercially synthesized in pUC57_Kan_gw_OK2 vector by Genewiz/Azenta. The sequence comprising a cassette that includes attP-FRT-splice acceptor-T2A-GAL4-poly(A)-3×P3EGFP-poly(A)-FRT-attP was transferred from the pM37 vector to the intermediary vector designed for homology, thereby generating a new homology donor vector. The construct was injected at 250 ng/µl final concentration into 664 embryos from the line: *y^1^w^∗^; attP40(y+){nos-Cas9(v+)}; iso50.* Eclosed flies were crossed with *y^1^ w\** flies, and the 3×P3–EGFP-positive progeny were selected based on GFP expression in the adult eyes.

### Protein Quantification

To determine human AXIN2 protein level upon overexpression in flies, *AXIN2* reference and variants were expressed pan-neuronally with *elav-GAL4*. Five fly heads from each genotype were homogenized in a cell lysis buffer (RIPA buffer, 1X liquid protease inhibitor (Gen DEPOT)) with an electric Dounce on ice. Next, Laemmli buffer and 5% β-mercaptoethanol (BME) was added to each tube and lysed heads were incubated on ice for 10 minutes and boiled for five minutes at 95°C. Samples were centrifuged at 14,000 RPM for five minutes at 4°C. The supernatant was loaded into 4-20% gradient gels (Mini-PROTEAN® TGX™), separated by SDS-PAGE, and transferred to nitrocellulose membranes (Bio-Rad). The following primary antibodies were used in this study: anti-Actin (mouse monoclonal: C4; 1:20,000) (EMD Millipore), anti-AXIN2 (rabbit monoclonal: 76G6; #2151; 1:1000) (Cell Signaling). Corresponding HRP-conjugated secondaries were used at 1:10,000 dilution.

### Immunostaining and imaging

The methods for preparing and visualizing *Drosophila* wing discs were performed as previously reported^60^. To summarize, wing discs from third-instar larvae at the wandering stage were dissected and preserved in 4% PFA. These samples were subsequently immunostained with the following antibodies: mouse anti-Wingless at a dilution of 1:50 (Developmental Studies Hybridoma Bank [DSHB], clone 4D4), mouse anti-Cut at the same dilution (DSHB, clone 2B10), and guinea pig anti-Senseless at a dilution of 1:100^61^. For the mounting phase, Vectashield (Vector Labs, H-1000-10) was employed. The wing discs were examined using a Zeiss LSM 880 laser confocal microscope. For the post-scanning image enhancement, ZEN software from Zeiss and Imaris software from Oxford Instruments were utilized.

### Adult wing and eye preparation and imaging

Adult *Drosophila* wings were dissected and mounted as previously described^60^. Briefly, wings from adult flies were excised and preserved in 70% ethanol before being arranged in a solution made from a mixture of CMCP-10 mounting medium (sourced from Masters in Wood Dale, IL) and lactic acid in a ratio of 3 to 1. For eyes, whole flies of appropriate genotypes were frozen at -20°C for 24 hours before imaging. The images for adult wings and eyes were captured using a Leica MZ16 stereomicroscope that was outfitted with an Optronics MicroFire camera and operated using Image-Pro Plus software version 7.0.

### Statistical Analyses

A two-sided Fisher’s exact test to determine if there was a significant difference between genotype distribution across E18.5 and postnatal timepoints for the *Axin2^E66K/+^* embryos and *Axin2^+/+^* littermates was done using GraphPad Prism (version 10.3.1 (509)). For *Drosophila* studies and quantification, ANOVA was followed by Tukey’s post hoc test using GraphPad Prism.

## Results

### *De novo* heterozygous variants clustered in the TNKS-binding domain of AXIN2 cause multiple congenital anomalies

A cohort consisting of three individuals with the same heterozygous *de novo* variant in *AXIN2* (NM_004655.4:c.196G>A, p.E66K) and overlapping phenotype consisting of features of an ectodermal dysplasia as well as skeletal and genitourinary anomalies was identified through the UDN and ClinVar. This variant was not present in gnomAD(v4.0.0)^41^ or in the BCM clinical site internal sequencing database. The CADD score is 27.2^62^, Revel score is 0.549^63^, and AlphaMissense score is 0.765^64^. The glutamic acid at this position is in the TNKS-binding domain of AXIN2^65^ and is conserved in vertebrates including mice and in zebrafish but not in *Drosophila melanogaster* (**Figure 1A and Figure S3A**). This variant has been reported in the Catalogue of Somatic Mutations in Cancer (COSMIC, v100) one time in a large intestinal adenocarcinoma from a 64-year-old male^66^.

**Figure 1.**
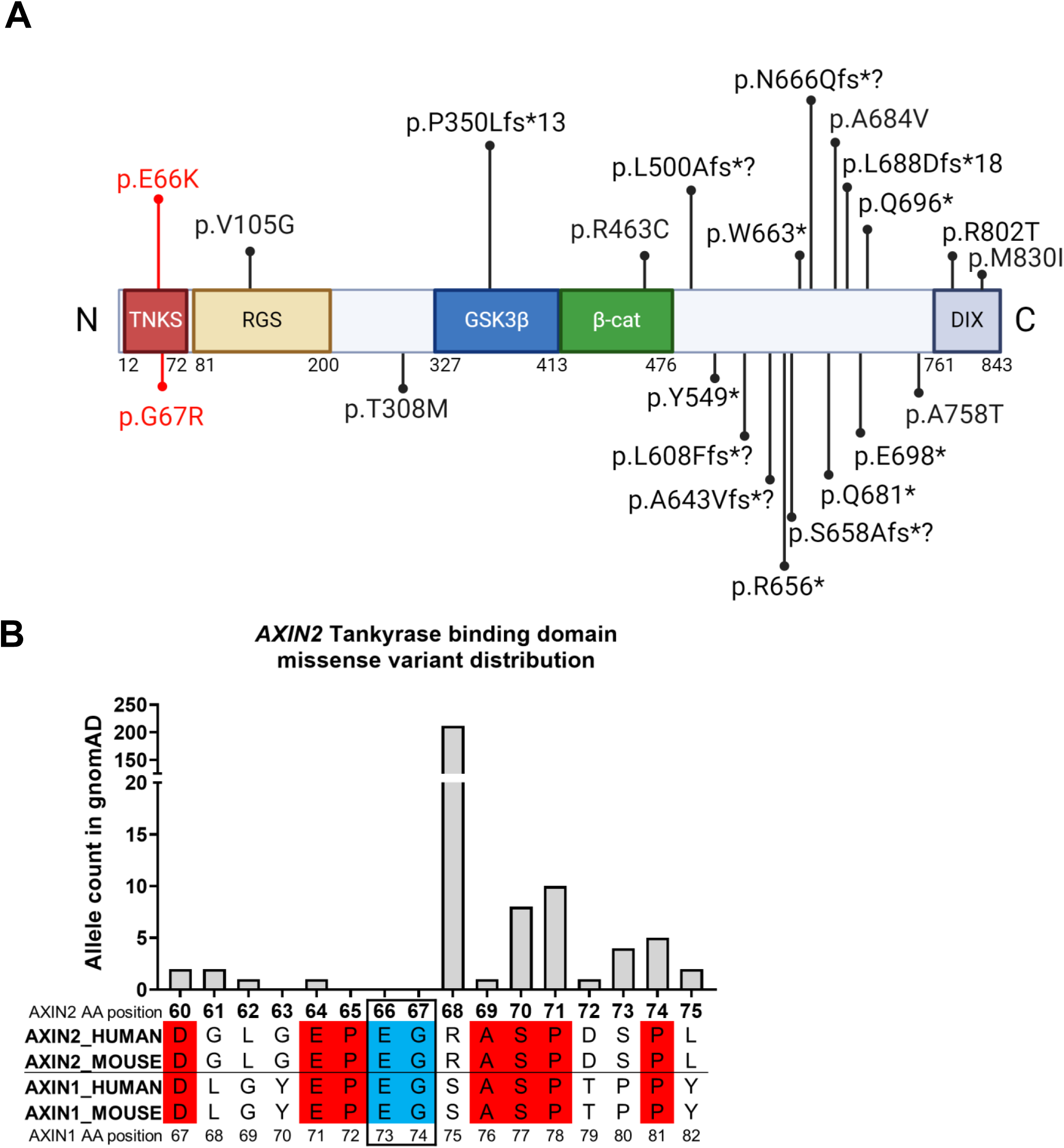
Participant Phenotypes. A) Pathogenic germline variants identified in *AXIN2*. The variants we identified are shown in red, previously published variants associated with oligodontia, oligodontia colorectal cancer syndrome, colon cancer and colon polyps previously published are shown in black^1-4,15,16^. TNKS = Tankyrase binding domain, RGS = Regulator of G-protein Signaling domain, GSK3β = Glycogen synthase kinase-3 beta binding domain, β-CAT = Beta-catenin binding domain, DIX = Disheveled and AXIN (DIX) dimerization domain. B) *AXIN2* missense allele counts derived from gnomAD aligned to human and mouse AXIN2 and AXIN1 amino acid alignments. Amino acids highlighted in red are conserved between all aligned proteins, and positions 66 and 67 are highlighted in blue. The absence of variation in gnomAD at positions 65 through 67 suggests that these residues may be critical for proper function of AXIN2. Conservation alignment was created using default parameters in Clustal Omega^43^. Amino acids 60-75 from AXIN2 are shown, and amino acids 67-82 from AXIN1 are shown. Variant allele counts were compiled from gnomAD_v4.0.0_ENSG00000168646_2.

Based on structural modeling data of AXIN1 and tankyrase interactions^37^, the proline at position 65 and the glycine at position 67 in AXIN2 are critical for the interaction between AXIN2 and tankyrase^37^. Given that there is an absence of missense variation at these positions in gnomAD(v4.0.0) (**Figure 1B, Figure S3B**) in conjunction with the conservation data, we hypothesized that variants impacting positions 65 and 67 might cause phenotypes similar to those with variants at position 66. Thus, we searched ClinVar for missense variation impacting these neighboring amino acids. This search identified participant 4 with a similar phenotype who has a *de novo* heterozygous variant at position 67 (NM_004655.4:c.199G>A, p.G67R). The CADD score of the p.G67R variant is 27.4, REVEL score is 0.494, and AlphaMissense score is 0.971. This glycine is conserved in vertebrates including mice and zebrafish but not in *Drosophila melanogaster*. This variant was also observed as a confirmed somatic heterozygous variant in a large intestinal adenocarcinoma sample from a female of unknown age and a diffuse adenocarcinoma of the stomach from a 38-year-old female in the COSMIC database(v100).

The phenotype observed in the four individuals with *de novo* heterozygous variants impacting the glutamic acid at position 66 or the glycine at position 67 of AXIN2 is complex and variable. Common phenotypes include digit and limb anomalies (4/4), global developmental delay (3/4), microcephaly (3/4), ophthalmologic complications (3/4), genitourinary anomalies (3/4), unique facial features (3/4), and palate abnormalities (2/4). Features suggestive of an ectodermal dysplasia such as oligodontia (3/4), sparse hair (3/4), sparse eyebrows and eyelashes (3/4), and nail abnormalities (2/4) are common. These findings together with sagittal craniosynostosis suggested a diagnosis of cranioectodermal dysplasia in participant 3. Although three of the four patients are children with complex phenotypes identified by exome or genome sequencing, one individual is an adult whose variant in *AXIN2* was identified on a colorectal cancer panel. This adult has a milder, but overlapping phenotype which includes digit abnormalities, skeletal abnormalities, and history of vesicoureteral reflux in addition to a diagnosis of colon cancer with >25 colon polyps. Detailed case reports are provided in supplemental material.

### Structural modeling

Since no atomic structure is available for the TNKS/AXIN2 complex, we attempted structure prediction of this complex *in silico* using AlphaFold2-multimer^67^ initially, and later using AlphaFold3^68^. TNKS proteins contain an ankyrin-repeat domain that consists of a series of ankyrin-repeat clusters (ARCs)^37^. In the top five prediction results from AlphaFold2-multimer, the first TNKS binding motif (TBM1) in AXIN2 binds the ankyrin-repeat domain ARC4 in all five predicted structures, while the second binding motif (TBM2) binds to ARC5 in one predicted structure. However, all five top predictions from AlphaFold3 shows only TBM2 binding to ARC5. Pairwise comparison of the Alphafold predicted structure and the known crystal structure of the ARC domain of TNKS with bound peptide (PDB IDs: 3UTM and 3TWW) shows that the peptide binding interactions are very similar, with the peptides interacting on the same surface of ARC. Currently, it is unclear which ARC of TNKS is the binding partner for each TBM. However, based on results from the pulldown assay and sequence conservation of critical residues in ARC implicated in TBM binding^37^, it is likely to be one of ARC2, ARC4, or ARC5.

The amino acid residues impacted by our variants are located in TBM2. Due to the ambiguity regarding which ARC is responsible for TBM2 binding and the relatively low confidence in the AlphaFold prediction of complex formation partners, we used the known crystal structure of the TNKS/AXIN1 complex^37^, in which each ARC2 of TNKS binds TBM1 and TBM2 of AXIN1, respectively, as a template for our *in silico* modeling of the variant structure. The arginine side chain of the mutated Gly67 was modeled using Coot. However, every rotamer conformation of the arginine clashes with surrounding TNKS residues in the complex, as there is no space to accommodate the bulky side chain (**Figure 2C**). As previously suggested by Morrone et al., the glycine-selection gate of TNKS is crucial for binding^37^. Consequently, the large side chain of the p.G67R variant of AXIN2 would prevent AXIN2 from binding.

**Figure 2.**
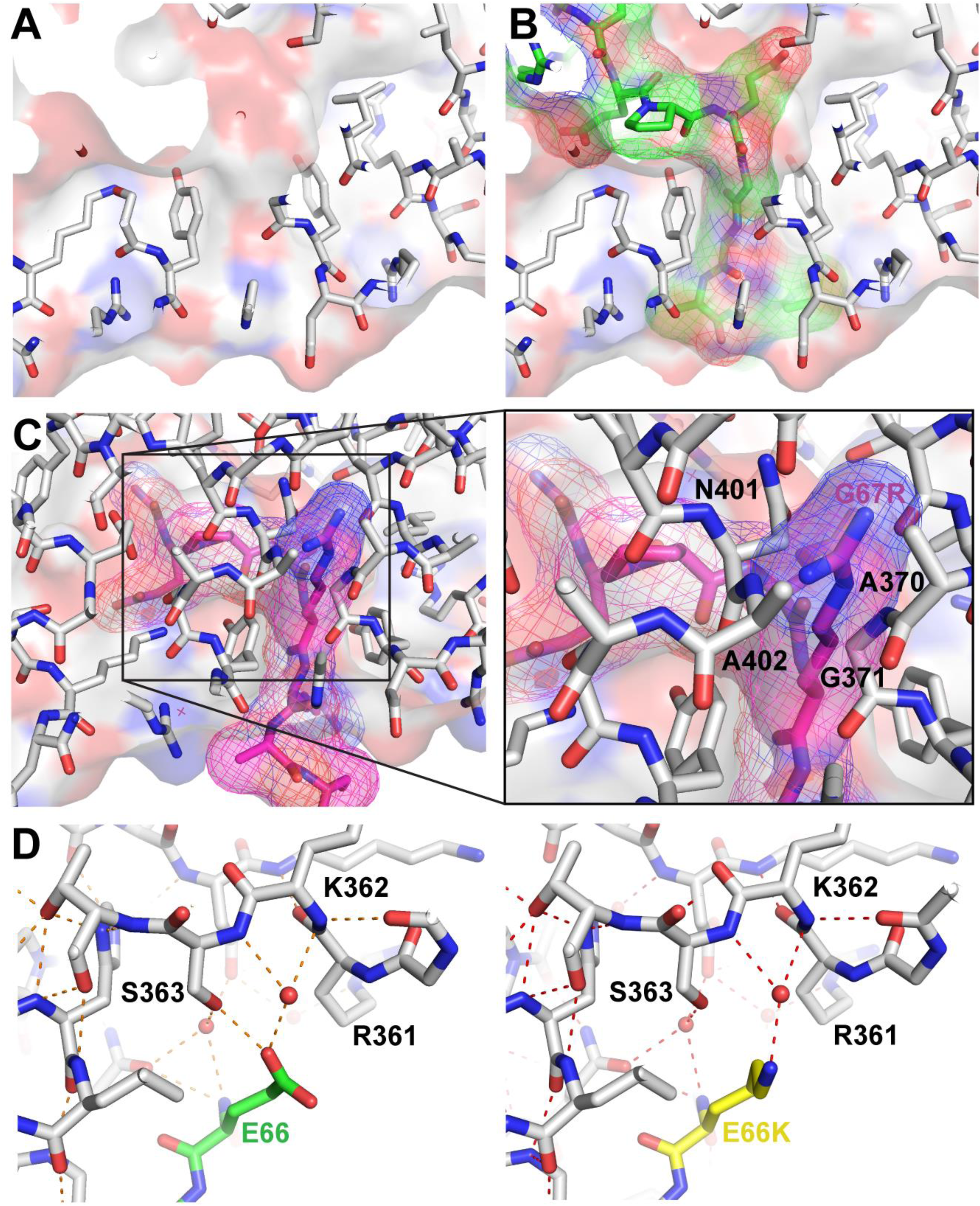
Structural Model of the AXIN2 variants. A) ARC2 of TNKS is shown as a stick model and surface representation. View looking from inside of the molecule toward the surface. B) Bound TBM2 of AXIN2 is shown in stick and surface representation and is colored in green. C) Bound TBM2 of the AXIN2 variant is shown in magenta. Enlarged view of the p.G67R mutant showing steric clashes with surrounding residues. D) The Glu66 side chain of AXIN2 TBM2 (green) participates in an extensive hydrogen bond network (dotted red line) that stabilizes complex formation. This hydrogen bond network would be disrupted by p.E66K mutation (yellow). Water molecules are shown in red balls. The model was generated based on the crystal structure of AXIN1 bound to TNKS (PDB ID: 3UTM).

Figure 2D shows the Glu66 side chain forming a hydrogen bond with the Ser363 side chain and water-mediated hydrogen bonds with the main chain N-H of K362 and S363, which participate in an extensive hydrogen bond network. The same hydrogen bond network, including water mediated hydrogen bond interactions, was also observed in the crystal structure of ARC4 of TNKS bound to a chimeric substrate peptide (PDB ID: 3TWW)^69^. These interactions would become less favorable when the glutamic acid is mutated to lysine due to its positive charge. The p.E66K side chain mutation model was generated using Coot, with a rotamer conformation that best maintained the hydrogen bond interactions. Currently, none of the versions of AlphaFold can predict water mediated interactions; therefore, relevant crystal structures were used as templates to analyze the mutant structure ^37,69^.

### Congenital anomalies in a p.E66K founder mouse embryo screen

We next used CRISPR/Cas9 genome editing in mice to confirm the pathogenicity and associated phenotypes of the p.E66K variant. Because we anticipated that the phenotypes associated with heterozygosity for p.E66K in humans would lead to postnatal lethality in mice, we employed a founder embryo screen (**Figure 3A**). CRISPR/Cas9-mediated HDR was used to generate embryos harboring an allele with the p.E66K variant and a neighboring synonymous variant (see Materials and Methods; **Figure S1**). At E18.5, embryos (n=37) were collected and each screened via Sanger sequencing and Synthego ICE analysis for the contribution of wild-type, indel, and p.E66K knock-in alleles (**Table S5**). Of the 37 embryos screened, 5 were wild-type and 26 harbored undesired indel mutations with or without the p.E66K knock-in allele. The remaining 6 embryos harbored only the p.E66K knock-in allele; however, only 2 of the 6 had knock-in allele contribution at or above 50% (96% and 53% knock-in allele contribution; sense and antisense ssODN targeting, respectively). Because low mosaicism for the knock-in allele may not be sufficient to produce phenotypes, μCT imaging was used to phenotype only these 2 knock-in embryos as well as a wild-type control sibling. Both p.E66K knock-in embryos displayed kinked tail and shortened soft palate abnormalities not observed in the wild-type control (Figure 3B-D).

**Figure 3.**
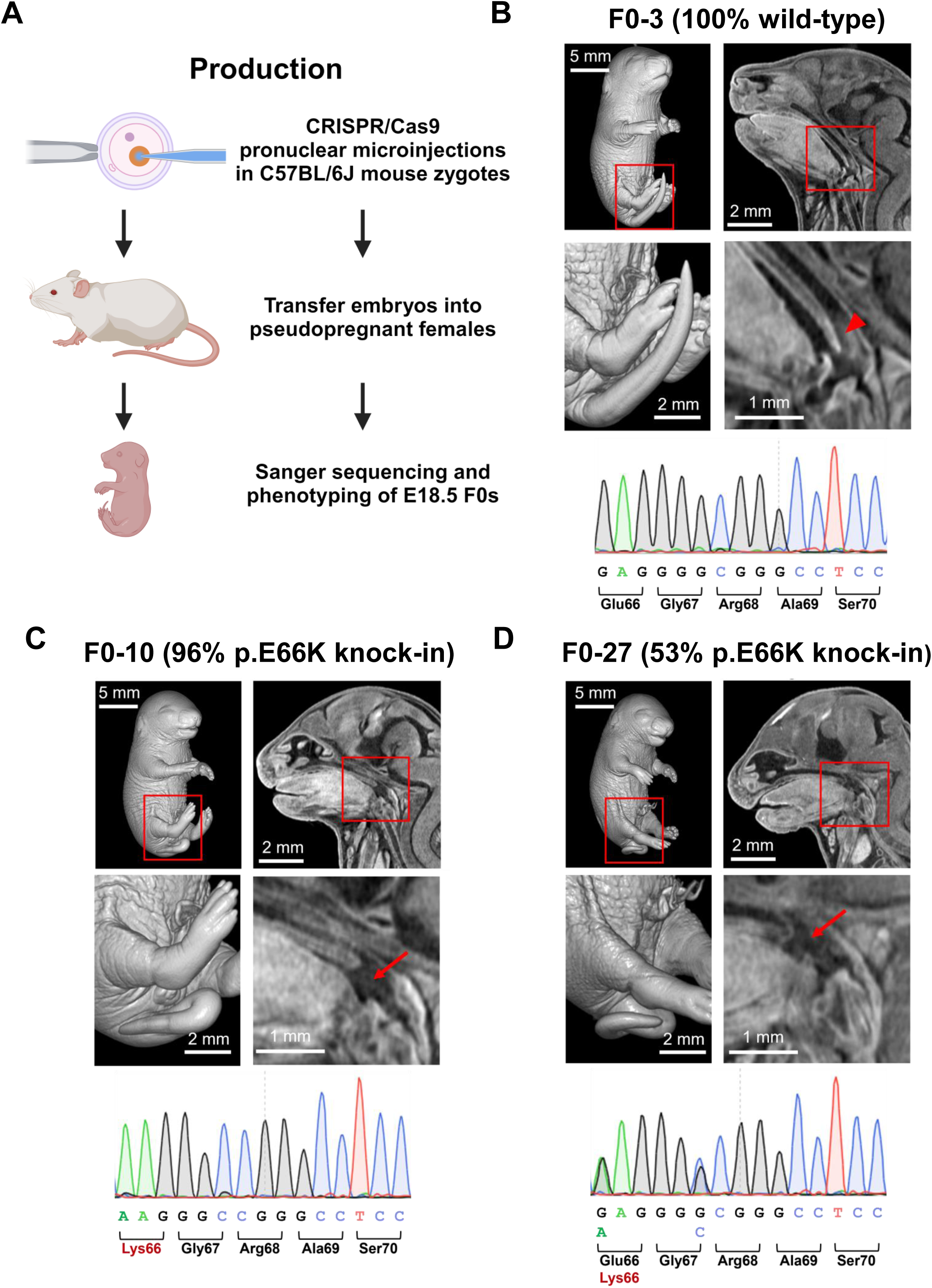
Production, genotyping, and phenotyping of p.E66K founder embryos. A) Outline of the process for generating and screening CRISPR founder embryos. B-D) Internal and external µCT images of embryos at E18.5 with respective Sanger sequence traces. B) A representative wild type embryo from the founder screen. No external anomalies are present (left images), and the soft palate is the expected length (right images). C) Embryo with 96% p.E66K knock-in allele contribution. This embryo had a kinked tail (left images) and had a shortened soft palate not extending to the epiglottis (right images). D) Embryo with 53% p.E66K knock-in allele contribution. This embryo also has a kinked tail (left images) and a shortened soft palate (right images). Bottom images are magnified views of the areas in red boxes on the top images. Arrowhead indicates normal soft palate extending to the epiglottis. Arrows indicate shortened soft palates.

### Confirmation of congenital anomalies using a p.E66K N1 mouse embryo screen

Although CRISPR founder screens bypass the need to generate a mouse line for phenotyping, mosaicism for the desired variant knock-in (HDR) allele, undesired indel (NHEJ) alleles, or both, in founders can confound detection of genotype-phenotype relationships^70^. Thus, we sought a mid-throughput approach to generate p.E66K heterozygous embryos for screening. We hypothesized that a CRISPR founder animal with low mosaicism for the p.E66K allele may survive to breeding age. From such an animal, N1 embryos heterozygous for the p.E66K allele could be consistently generated, and genotype-phenotype associations could be more easily established. To produce such a founder, we turned to prime editing, which has been shown to be inefficient when editing in the mouse germline^71-74^.

Using prime editing, we produced a viable male founder that was 20% mosaic for an allele harboring the p.E66K variant and two neighboring synonymous variants (*Axin2^E66K^*) (see Materials and Methods; **Figure S2**). Matings using the male founder produced six litters with a total of 30 liveborn N1 offspring, which were genotyped at 2-3 weeks of age. Interestingly, no heterozygous *Axin2^E66K/+^* offspring were identified (**Figure 4B**). Due to these results, we hypothesized that there was either no germline transmission of the p.E66K variant or the variant was associated with pre-weaning lethality. Therefore, we performed *in vitro* fertilization with founder sperm to determine if *Axin2^E66K/+^* N1 embryos were present and viable just prior to birth (E18.5). Of 29 viable N1 embryos collected at E18.5, 14 (48%) were heterozygous for the p.E66K variant allele. A Fisher’s exact test comparing genotypes detected at E18.5 and weaning age showed a significant difference (p<0.0001) (**Figure 4B**). External examination of the 14 *Axin2^E66K/+^* E18.5 embryos revealed kinked tail abnormalities (N=5; 36%), missing digit (N=1; 7%), and edema or hydrocephalus (N=2; 14%). These phenotypes were not observed in 9 *Axin2^+/+^* littermate controls. We performed μCT imaging on the *Axin2^+/+^* and *Axin2^E66K/+^* embryos, which confirmed the observed external phenotypes (**Figure 4C&D; Figure S4**). Critically, μCT imaging also revealed a shortened soft palate phenotype in all *Axin2^E66K/+^* embryos, which was not observed in any *Axin2^+/+^* controls (**Figure 4D; Figure S5**). Together, the results from our N1 embryo screen demonstrate that heterozygosity for *Axin2^E66K^* causes lethality between birth and weaning and embryonic phenotypes like those observed in the founder screen and in the participants. Moreover, these results suggest that lethality may be secondary to the shortened palate observed in *Axin2^E66K/+^*embryos.

**Figure 4.**
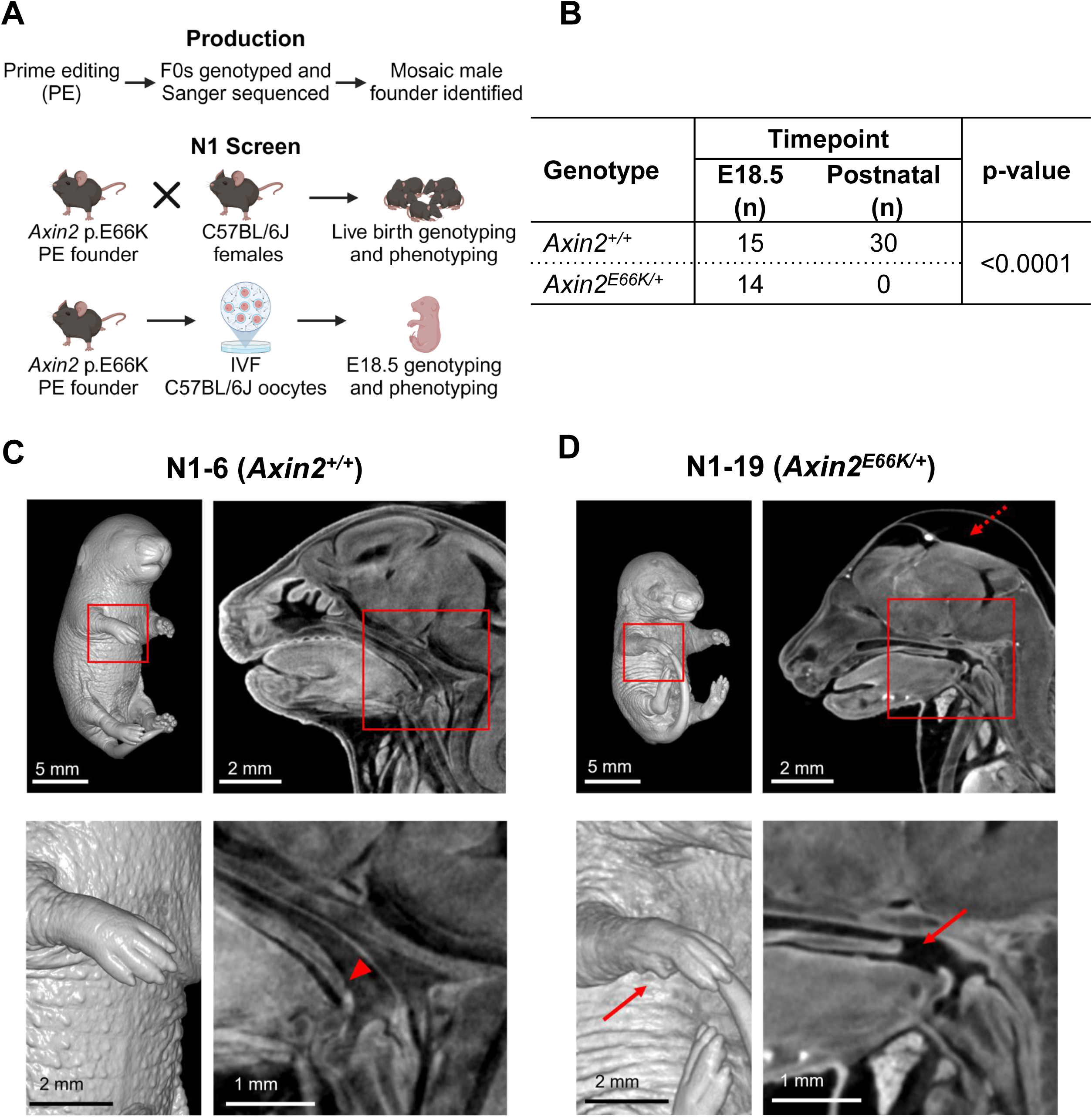
Production, genotyping, and phenotyping of the p.E66K knock-in mouse model. A) Outline of the mosaic founder production and N1 screen for the p.E66K knock-in allele generated with prime editing. B) Genotype distribution of N1 E18.5 embryos and liveborn animals generated from the p.E66K mosaic founder. There were no N1 *Axin2^E66K/+^* animals at weaning age, whereas about 50% of the embryos at E18.5 were carriers (p < 0.0001, Fisher’s exact for genotype distribution across timepoints). C) A representative E18.5 wild type embryo from the N1 screen. No external anomalies are present (left images), and the soft palate is the expected length, extending to the epiglottis (right images). D) A representative E18.5 *Axin2^E66K/+^* embryo with missing forelimb 5^th^ digit (left images) and edema and a shortened soft palate not extending to the epiglottis (right images). Bottom images are magnified views of the areas in red boxes on the top images. Dashed arrow indicates edema. Arrowhead indicates normal soft palate extending to the epiglottis. Arrows indicate missing digit and shortened soft palate.

### The p.E66K variant allele functions differently than *AXIN2* reference and pathogenic ODCRCS variant alleles in *Drosophila*

Although the Glu66 residue is not conserved in *Drosophila melanogaster,* flies remain an important *in vivo* system for assessing variant function. Hence, we generated transgenic flies that express AXIN2 reference (*AXIN2^REF^*) or variants under the control of an Upstream Activation Sequence (UAS). We generated the p.E66K variant (*AXIN2^E66K^*) present in three of the participants as well as two pathogenic truncating variants (*AXIN2^R656X^* and *AXIN2^W663X^*) observed in oligodontia-colorectal cancer syndrome (ODCRCS)^3,9^. All transgenes were inserted in the same genomic site in the flies, controlling for positional effects, and ensuring similar protein expression levels (**Figure S6A**). In parallel, we generated an *Axn^TG4^* (*Axn^T2A-GAL4^*) mutant *Drosophila* allele as part of a large-scale resource^59^. As expected, this allele was homozygous lethal and remains lethal when *in trans* with a corresponding deficiency line (*Df(3R)X3F*) lacking the *Axn* gene, indicating the *Axn^TG4^*mutant is loss-of-function. The *Axn^TG4^* allele expresses a GAL4 transcriptional activator in the same spatial and temporal pattern as the *Drosophila* gene, *Axn*. We found that expressing human *AXIN2^REF^* in heterozygous *Axn^TG4^*flies (*Axn^TG4/+^* > *UAS-AXIN2^REF^/+*) led to no adult flies at 18°C and 22°C. These data suggested that *Axn*/*AXIN2* is likely dosage-sensitive in flies which prevented us from conducting humanization-based rescue studies.

As an alternative to humanization, we turned to an overexpression-based strategy for assessing the impact of reference versus variant human cDNA expression in flies in a tissue-specific manner^75^. Given that AXIN2 has a known role in β-catenin-dependent Wnt signaling^17^, we opted to overexpress *AXIN2^REF^*and variants in the developing wing of the fly using *nubbin-GAL4*^76^ where Wnt (wingless in flies) signaling is well characterized. The expression of *AXIN2^REF^* caused wing margin loss marked by posterior serration and bristle loss (**Figure 5A**). This phenotype was enhanced by increasing the temperature as GAL4-mediated UAS expression of the transgene is positively correlated with temperature^77^ (**Figure 5A**; **Figure S6B**). Importantly, overexpression of a GFP-tagged fly Axn (*Axn^GFP^*)^18^ causes a similar phenotype, albeit more severe. These data indicate that Axn and AXIN2 exert similar effects in the developing wing disc. In this context, the *AXIN2^E66K^* variant leads to a consistently worse phenotype than *AXIN2^REF^* as demonstrated by smaller wings as well as the presence of ectopic bristles and blisters (**Figure 5A**; **Figure S6B**). These data indicate that the p.E66K may act as a gain-of-function with potential neomorphic functions. The *AXIN2^R656X^*and *AXIN2^W663X^* variants observed in ODCRCS caused an even stronger phenotype than both *AXIN2^REF^* or *AXIN2^E66K^*as wings were smaller with more pronounced wing margin loss across temperatures. These data indicate that the ODCRCS variants may also act as a gain-of-function.

**Figure 5.**
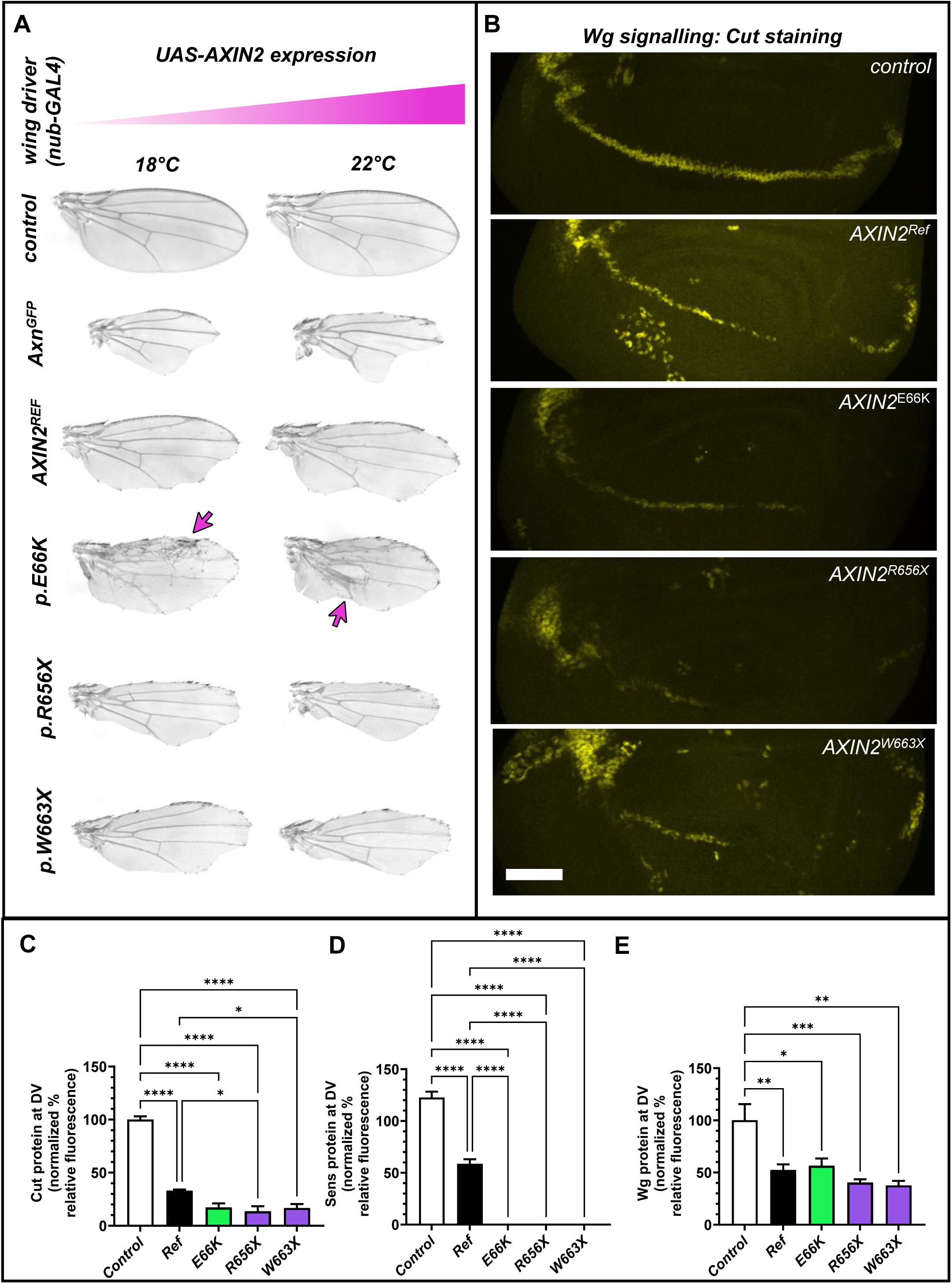
The p.E66K acts as a gain-of-function variant in the *Drosophila* wing. A) Representative images of adult fly wings expressing UAS-cDNA using *nubbin-GAL4* at the temperatures indicated. A minimum of 60 wings were examined from three independent crosses. Phenotypes were nearly 100% penetrant. B) Representative immunostaining of the Cut protein in the developing wing disc at 22°C upon UAS-cDNA overexpression with *nubbin-GAL4*. Scale bar = 40 μm. C-E) Quantification of fluorescence signal for Cut, Sens, and Wingless (Wg) proteins in the developing wing disc upon overexpression of UAS-cDNA using *nubbin-GAL4* at 22°C. Immunostaining images for Sens and Wg are provided in Figure S7. ANOVA followed by Tukey’s post hoc test. *p<0.05, **p<0.01, ***p<0.001, ****p<0.0001.

To confirm that *AXIN2* overexpression impacts wingless and that downstream canonical signaling (Wnt/β-catenin signaling) is affected at the cellular level, we examined the developing wing disc in third instar larvae. Compared to control wing discs expressing lacZ, the expression of *AXIN2^REF^* caused a decrease in Cut staining, a Wingless downstream effector (**Figure 5B**). This diminished Cut expression is further reduced when the *AXIN2^E66K^* or the ODCRCS variants are overexpressed in the developing wing disc, but only statistically significant with the ODCRCS variants suggesting an intermediate function of the p.E66K variant (**Figure 5C**). In line with this, the expression of Sens, a wingless responsive transcription factor expressed adjacent to wingless secreting cells of the dorsal-ventral boundary is decreased in a similar fashion (**Figure S7A)**. Sens expression is almost completely absent when either the *AXIN2^E66K^* or the ODCRCS variants are overexpressed in the developing wing disc (**Figure 5D**). Intriguingly, the Wingless ligand is also diminished in *AXIN2^REF^, AXIN2^E66K^* and the ODCRCS variants to similar degrees (**Figure 5E and Figure S7B).** These data indicate that both the p.E66K and truncations observed in ODCRCS inhibit wingless signaling (WNT signaling) in this tissue and can act as gain-of-function variants in this context.

To examine the impact of *AXIN2* variants in another cell context, we chose to overexpress *AXIN2* in the developing *Drosophila* eye using *ey-GAL4*. Although typically a dispensable tissue in flies, expression with *ey-GAL4* can sometimes lead to lethality. Here, overexpression of fly *Axn^GFP^* with *ey-GAL4* causes 100% pupal lethality at 22°C. Overexpression of *AXIN2^REF^*or *AXIN2^E66K^* causes ∼75% and ∼50% pupal lethality, respectively (**Figure 6A**). In surviving progeny, flies expressing *AXIN2^REF^* display a smaller eye compared to *AXIN2^E66K^* (**Figure 6B-C**). These data suggest that when expressed in the fly eye, the p.E66K acts as a loss-of-function. On the contrary, the *AXIN2^R656X^* and *AXIN2^W663X^* variants observed in ODCRCS cause almost ∼100% pupal lethality with the few survivors displaying extremely small eyes (**Figure 6C**). Hence, like their function in the wing, the truncations observed in ODCRCS consistently display gain-of-function activity.

**Figure 6.**
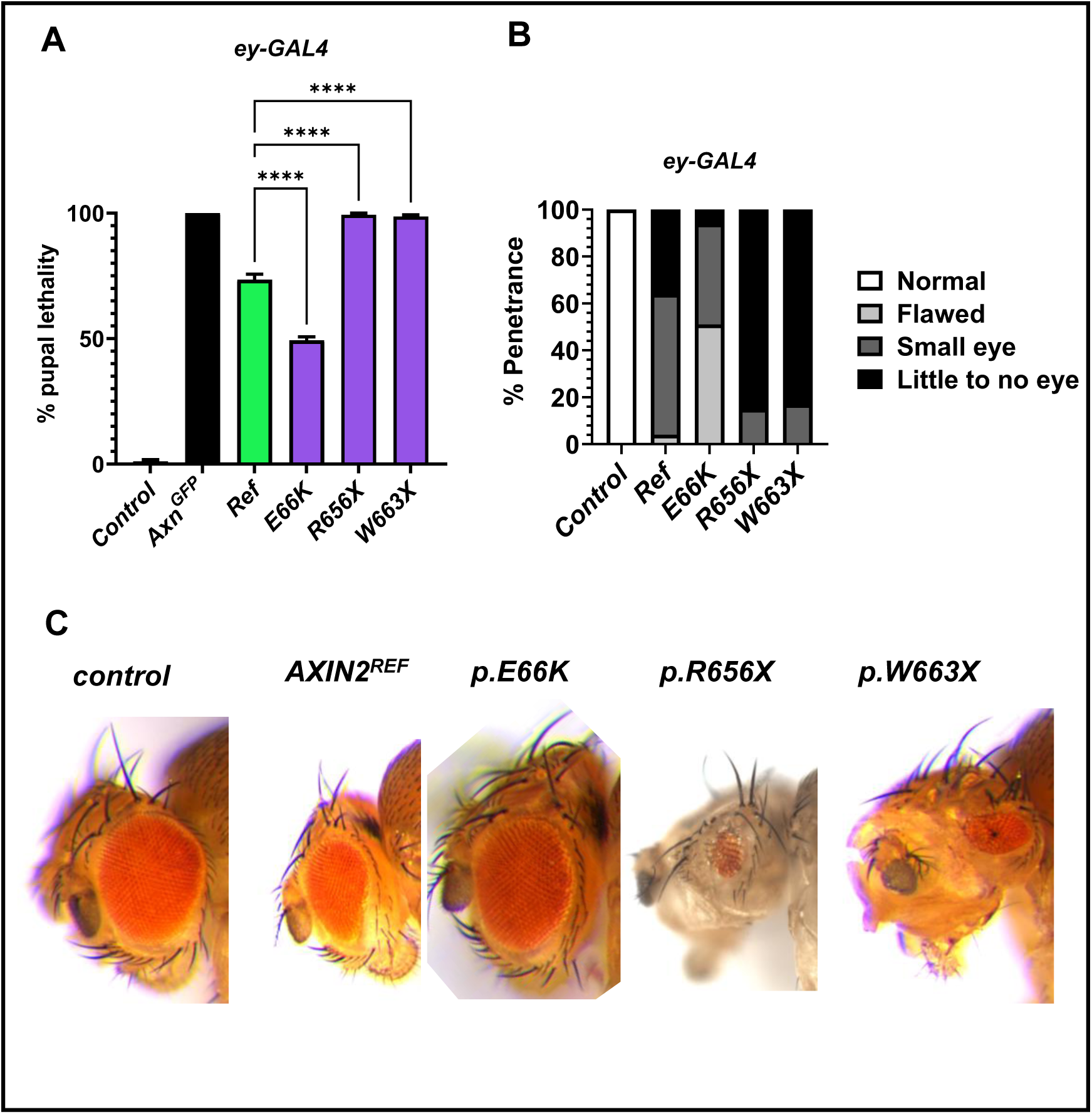
The p.E66K acts as a loss-of-function variant in the *Drosophila* eye. A) Quantification of pupal lethality upon overexpression of UAS-cDNA with ey-GAL4 at 22°C. ANOVA followed by Tukey’s post hoc test. ****p<0.0001. B) Range of eye phenotypes identified upon overexpression of UAS-cDNA with *ey-GAL4* at 22°C in surviving adult flies. C) Representative images of eye phenotypes upon overexpression of UAS-cDNA with *ey-GAL4* at 22°C in surviving adult flies.

## Discussion

In this study, we identified four individuals who had previously unreported germline variants (p.E66K and p.G67R) in the TNKS-binding domain of *AXIN2* and a phenotype that only partially overlaps with ODCRCS, the only known Mendelian disorder associated with heterozygous pathogenic variants in this gene^1-4,9,15,16^. Published phenotypes associated with germline *AXIN2* variants include sparse hair and eyebrows, colonic polyps/polyposis, colorectal cancer, and oligodontia or tooth agenesis^1-4,9,15,16,34^. Two recent publications suggested that cleft lip/palate, olfactory neuroblastoma, and gastric adenoma should be included in the spectrum of germline *AXIN2-*associated phenotypes^78,79^. In addition to several common ODCRCS phenotypes, the patients in our cohort share several other overlapping phenotypes that include developmental delays, microcephaly, digit and limb anomalies, ophthalmologic complications, genitourinary anomalies, and unique facial features. Thus, the phenotypes caused by p.E66K and p.G67R variants in *AXIN2* expand the phenotypic spectrum of AXIN2-related disorders. Pathogenic variants in *AXIN2* associated with ODCRCS have been reported to impact multiple domains except for the TNKS-binding domain^1-4,9,15,16,34,78^. To our knowledge, these two variants described here are the first *AXIN2* pathogenic variants that impact the TNKS-binding domain.

Relative to ODCRCS, the more severe phenotypes of our patients with p.E66K or p.G67R variants suggest the variants in the TNKS-binding domain of AXIN2 have a unique impact on AXIN2 function. TNKS poly-ADP-ribosylates AXIN2, which leads to the ubiquitination and proteasomal degradation of AXIN2^35,37^. Our structural modeling studies suggest that these variants may directly impact TNKS binding to AXIN2. In particular, the p.G67R variant is predicted to completely abolish AXIN2 binding to TNKS, while the p.E66K variant is predicted to weaken the interaction with the main chain N-H of K362/S363 due to a less favorable charge distribution, although a polar interaction with Serine 363 is still possible. In vitro studies of a p.G74V mutation in AXIN1 (which is equivalent to p.G67V in AXIN2) demonstrated that this mutation prevents TNKS from binding to AXIN1 in GST-pulldown assays, a finding that supports our data^37^. Thus, a p.E66K or p.G67R variant may prevent TNKS and AXIN2 binding, which may lead to increased stability of AXIN2.

The modeling of *de novo* heterozygous variants associated with severe early-onset phenotypes is challenging in mice using traditional constitutive knock-in strategies due to expected lethality. Thus, to circumvent this challenge, we used two mouse embryo screening approaches to confirm the pathogenicity and phenotypes of the p.E66K variant, which was identified in 3 of the 4 participants in our cohort. Using both a CRISPR founder embryo screen and an innovative prime editing N1 embryo screen, the p.E66K variant was found to cause shortened soft palate, kinked tail, and digit abnormalities, phenotypes resembling the palate and skeletal abnormalities observed in the participants. Moreover, the observed perinatal lethality of N1 mice heterozygous for the p.E66K variant further supported pathogenicity. Given that the soft palate elevates during swallowing to prevent food and fluids from going into the nasopharynx, we propose that p.E66K heterozygous pups die shortly after birth due to nasal regurgitation. Importantly, the phenotypes observed in mouse embryos heterozygous for the p.E66K variant contrast with the those in published reports describing *Axin2* homozygous knockout mice, which are viable, fertile, and have abnormal cranial suture closure^13,80-82^. This suggests that the effect of the p.E66K variant on AXIN2 function is not simply loss-of-function.

Our fly modeling supports a complex and cell context-dependent effect of the p.E66K mutation on AXIN2 function. Overexpression of the p.E66K and ODCRCS variant alleles caused more severe phenotypes than overexpression of reference AXIN2 in the fly wing, indicating gain-of-function for all variants tested. Overexpression of the p.E66K variant in the developing fly wing also caused ectopic bristles, which were not observed in reference or ODCRCS variant expressing wings, indicating potential neomorphic activity. In contrast, overexpression of the p.E66K variant in the developing *Drosophila* eye caused less severe phenotypes compared to reference, which is consistent with AXIN2 loss-of-function. Critically, overexpression of the ODCRCS variants in the eye produced phenotypes consistent with AXIN2 gain-of-function, like their overexpression in the wing. Thus, the effect of the p.E66K variant on AXIN2 function is complex, context-dependent, and different than the ODCRCS variants, revealing a potential underlying mechanism for phenotypic expansion.

*In vitro*, many ODCRCS variants produce premature stop codons distributed throughout *AXIN2*, but some of these mutant transcripts appear to escape nonsense-mediated decay^83^. Consequently, truncated AXIN2 protein is produced, retains AXIN2 function, and demonstrates increased stability in some situations^83^. These molecular characteristics are consistent with the gain-of-function phenotypes observed when these variants are overexpressed in both the fly wing and eye. Why the p.E66K variant in the TNKS binding domain revealed differential, cell context-specific effects on AXIN2 function (i.e., gain-of-function versus loss-of-function) in our fly models remains to be resolved. Cell context-specific roles for TNKS in the regulation of AXIN2 protein levels could play a role.

The function of AXIN2 in canonical Wnt signaling is primarily considered to be inhibitory, as AXIN2 is a critical component of the β-catenin destruction complex in the absence of Wnt signaling. However, once Wnt signaling is active, AXIN2 binds to and plays an important role in maintaining the phosphorylation of the Lrp5/6 receptors, effectively promoting Wnt signaling^29-32^. The phenotypes observed in our mouse and fly models support this dichotomous role for AXIN2 in canonical Wnt signaling, which may also be cell context-dependent. Canonical Wnt/β-catenin signaling in cranial neural crest (CNC)-derived mesenchymal cells is essential for proper development of the soft palate, and the shortened soft palate phenotype in p.E66K heterozygous embryos is consistent with disruption of Wnt signaling in these cells^84,85^. On the other hand, the kinked tail phenotype observed in p.E66K heterozygous embryos is consistent with hyperactivation of Wnt signaling in the neural tube^86^. Interestingly, previous studies of mouse embryos harboring an ENU-induced p.V26D mutation in the tankyrase binding domain of *Axin2* (*Axin2^canp^*, **Figure S3**) revealed similar cell context-dependent effects with inhibition of Wnt signaling in many embryonic tissues but a gain of Wnt signaling in the primitive streak^36^. Similarly, the phenotypes caused by AXIN2 overexpressed in the *Drosophila* wing are consistent with inhibition of Wnt signaling^61,87^; the phenotypes induced by AXIN2 overexpressed in the *Drosophila* eye are suggestive of Wnt signaling activation as Wnt plays an inhibitory role in Drosophila eye development^88^.

Based on patient phenotypes and our animal modeling data, we propose that specific variants impacting the TNKS-binding domain of AXIN2 are associated with phenotypic expansion and a novel Mendelian disorder with an underlying complex molecular mechanism. Future studies will explore the cell context-specific effects of TNKS-binding domain variants on AXIN2 stability, AXIN2 function, and Wnt/β-catenin signaling. The modeling strategies used to demonstrate the pathogenicity of this variant may be beneficial for the resolution of other *de novo* heterozygous variants of uncertain significance associated with congenital anomalies in humans.

## Data Availability

All data produced in the present study are available upon reasonable request to the authors except if limited by stipulations by the Institutional Review Board.

## Acknowledgements

We thank the patients and families for participating in this study. Clinical research reported in this publication was supported in part by National Institutes of Health (NIH) U01HG007709 to BL and U01NS134348 to BL and JR. The genome sequencing for participant 1 was done through the UDN Clinical Sequencing Core supported by NIH U01HG007942. Use of the Baylor College of Medicine Intellectual and Developmental Disabilities Research Center Clinical translational core was supported by P50HD103555. Fly modeling was supported by NIH U54NS093793, R24OD022005, and R24OD031447 to HJB, OK and SY. Mouse modeling was supported by NIH U54OD030165 to JDH and LCB. Use of the Macromolecular X-ray Crystallography Core at Baylor College of Medicine was supported in part by NIH S10OD030246 to SL. Use of the Optical Imaging & Vital Microscopy Core at the Baylor College of Medicine and the MicroCT Imaging Facility at the McGovern Medical School at UTHealth was supported in part by NIH S10OD030336. Use of the BCM Genetically Engineered Rodent Models Core was supported in part by the Dan L. Duncan Comprehensive Cancer Center Support grant P30CA125123. The content is solely the responsibility of the authors and does not necessarily represent the official views of the National Institutes of Health. The mention of trade names, commercial products, or organizations do not imply endorsement by the US Government. LCB and HTC are supported by a Burroughs Wellcome Fund Career Award for Medical Scientists. HTC is supported by the Cain Pediatric Neurology Research Foundation Laboratories and McNair Medical Institute at The Robert and Janice McNair Foundation.

## Supplemental Material

### Supplemental Figures

**Figure S1. Genome editing approaches for the p.E66K founder embryos.** A) Genome editing with a sense strand derived sgRNA and anti-sense strand derived ssODN. B) Genome editing with a sense strand derived sgRNA and anti-sense strand derived ssODN. Scissors indicate sgRNA cut sites; asterisks indicate p.E66K mutation and p.G67 synonymous mutation. The location of primers (E2 F and E2 R) used for PCR genotyping and Sanger sequencing are shown.

**Figure S2. Production and characterization of *Axin2^E66K/+^* mosaic founder and N1 embryos.** A) Prime editing production of a mosaic founder. Orange scissors indicate the pegRNA; green scissors indicate the sgRNA for DNA nicking. The asterisk indicates p.E66K mutation and p.G67 synonymous mutation. Insertion of the p.E66K mutation generates a StyI restriction endonuclease cut site. The location of primers (E2 F and E2 R) used for PCR genotyping and Sanger sequencing are shown. The 653 bp wild type PCR product is not digested by StyI; the 653 bp p.E66K variant allele PCR products are digested by StyI, resulting in 335 and 318 bp products. B) Sanger sequencing traces of the mosaic founder, a representative Axin2^E66K/+^ N1 embryo, and a representative wild type N1 embryo. The founder male is mosaic for an allele harboring the p.E66K point mutation, a p.G67 synonymous mutation, and a p.S70 (TCG>TCC) synonymous mutation. Co-transmission of the variants to N1 offspring confirms linkage. C) Representative PCR-StyI restriction digest genotyping results for N1 embryos. NTC = no template control.

**Figure S3**. **Extended gnomAD conservation figure and *AXIN2* conservation across multiple species.** A) Conservation alignment between *Homo sapiens* AXIN2, *Drosophila melanogaster* AXN, and AXIN2 from various organisms (see methods and materials for full list). Amino acids highlighted in red are 90-100% identical in the alignment, orange is 80-89% identical, yellow is 70-79% identical. Positions 66 and 67 in human AXIN2 are highlighted in blue, p.E66 and p.G67 are conserved in all organisms aligned except *D. melanogaster*. B) *AXIN2* missense allele counts derived from gnomAD aligned to *Homo sapiens* and *Mus musculus* AXIN2, and *Homo sapiens* and *Mus musculus* AXIN1. Red color highlights amino acids conserved between all aligned proteins, positions 66 and 67 are highlighted in blue. The absence of variation in gnomAD at positions 65 through 67 suggests that these residues may be critical for proper function of AXIN2. Missense variant allele counts were compiled from gnomAD_v4.0.0_ENSG00000168646_2.

**Figure S4. External imaging of embryos from the N1 screen.** External μCT images of E18.5 wild type (WT, *Axin2^+/+^*) and p.E66K heterozygous (Het, *Axin2^E66K/+^*) embryos from the N1 screen. No external anomalies are present in wild type embryos; missing digit (N1-19) and kinked tail phenotypes (N1-11, 15, 16, 22, 23) are observed in *Axin2^E66K/+^* embryos (arrows).

**Figure S5. Internal imaging of embryos from the N1 screen.** Internal, sagittal μCT images of E18.5 wild type (*Axin2^+/+^*) and p.E66K heterozygous (*Axin2^E66K/+^*) embryos from the N1 screen. No internal anomalies are present in wild type embryos, with the soft palate extending to the epiglottis (box); all *Axin2^E66K/+^* embryos have a shortened soft palate not extending to the epiglottis (box).

**Figure S6. The p.E66K variant is stable and acts as a gain-of-function variant in the *Drosophila* wing.** A) Representative western blot showing expression of UAS-AXIN2 cDNA constructs are stable and relatively similar in level. B) Representative images of adult fly wings expressing UAS-cDNA using *nubbin-GAL4* at the temperatures indicated. A minimum of 60 wings were examined from three independent crosses. Phenotypes were nearly 100% penetrant.

**Figure S7. The p.E66K variant leads to increased inhibition of Wingless signaling.** A-B) Representative immunostaining of the Sens and Wg protein in the developing wing disc at 22°C upon UAS-cDNA overexpression with *nubbin-GAL4*. Scale bar = 40 μm.

### Supplemental Tables

**Table S1** – Guides for *Axin2* founder screen by traditional CRISPR HDR.

**Table S2** – T Guides for Axin2 mouse model production by prime editing.

**Table S3** – Prime editing pegRNA sequences.

**Table S4** – Primers used for site-directed mutagenesis of AXIN2 for fly modeling.

**Table S5** – Results of the ICE analysis of Sanger sequencing reads from p.E66K founder embryos.

## Member Lists

Members of the UDN and members of the BCM CPMM lists are provided in supplemental material.

### Web Resources

AlphaFold2 Multimer, https://github.com/sokrypton/ColabFold

AlphaFold3, https://alphafoldserver.com

CADD, https://cadd.gs.washington.edu/

Catalogue of Somatic Mutations in Cancer (COSMIC), https://cancer.sanger.ac.uk/cosmic

Clinvar, https://www.ncbi.nlm.nih.gov/clinvar/

Clustal Omega, http://www.clustal.org/

Codified Genomics, https://codifiedgenomics.com

Developmental Studies Hybridoma Bank, https://dshb.biology.uiowa.edu/

Gnomad, https://gnomad.broadinstitute.org/

OMIM, https://www.omim.org/

Mouse Genome Informatics (MGI), https://www.informatics.jax.org/

Pegfinder, http://pegfinder.sidichenlab.org

REVEL, https://sites.google.com/site/revelgenomics/

Wellcome Sanger Institute Genome Editing (WGE), https://wge.stemcell.sanger.ac.uk/

Synthego Inference of CRISPR Edits (ICE), https://ice.synthego.com/#/

## References

1. Beard, C., Purvis, R., Winship, I.M., Macrae, F.A., and Buchanan, D.D. (2019). Phenotypic confirmation of oligodontia, colorectal polyposis and cancer in a family carrying an exon 7 nonsense variant in the AXIN2 gene. Familial Cancer 18, 311–315. 10.1007/s10689-019-00120-0.

2. Jensen, J.M., Skakkebæk, A., Gaustadness, M., Sommerlund, M., Gjørup, H., Ljungmann, K., Lautrup, C.K., and Sunde, L. (2022). Familial colorectal cancer and tooth agenesis caused by an AXIN2 variant: how do we detect families with rare cancer predisposition syndromes? Familial Cancer 21, 325–332. 10.1007/s10689-021-00280-y.

3. Lammi, L., Arte, S., Somer, M., Järvinen, H., Lahermo, P., Thesleff, I., Pirinen, S., and Nieminen, P. (2004). Mutations in *AXIN2* Cause Familial Tooth Agenesis and Predispose to Colorectal Cancer. The American Journal of Human Genetics 74, 1043-1050. 10.1086/386293.

4. Yue, H., Liang, J., Yang, K., Hua, B., and Bian, Z. (2016). Functional analysis of a novel missense mutation in AXIN2 associated with non-syndromic tooth agenesis. European Journal of Oral Sciences 124, 228–233. 10.1111/eos.12273.

5. Arenas, E. (2014). Wnt signaling in midbrain dopaminergic neuron development and regenerative medicine for Parkinson’s disease. Journal of Molecular Cell Biology 6, 42–53. 10.1093/jmcb/mju001.

6. Jia, L., Piña-Crespo, J., and Li, Y. (2019). Restoring Wnt/β-catenin signaling is a promising therapeutic strategy for Alzheimer’s disease. Molecular Brain 12, 104. 10.1186/s13041-019-0525-5.

7. Wang, Q., Huang, X., Su, Y., Yin, G., Wang, S., Yu, B., Li, H., Qi, J., Chen, H., Zeng, W., et al. (2022). Activation of Wnt/β-catenin pathway mitigates blood–brain barrier dysfunction in Alzheimer’s disease. Brain 145, 4474–4488. 10.1093/brain/awac236.

8. Lancaster, M.A., Louie, C.M., Silhavy, J.L., Sintasath, L., DeCambre, M., Nigam, S.K., Willert, K., and Gleeson, J.G. (2009). Impaired Wnt–β-catenin signaling disrupts adult renal homeostasis and leads to cystic kidney ciliopathy. Nature Medicine 15, 1046–1054. 10.1038/nm.2010.

9. Marvin, M.L., Mazzoni, S.M., Herron, C.M., Edwards, S., Gruber, S.B., and Petty, E.M. (2011). AXIN2-associated autosomal dominant ectodermal dysplasia and neoplastic syndrome. American Journal of Medical Genetics Part A 155, 898–902. 10.1002/ajmg.a.33927.

10. Yook, J.I., Li, X.-Y., Ota, I., Hu, C., Kim, H.S., Kim, N.H., Cha, S.Y., Ryu, J.K., Choi, Y.J., Kim, J., et al. (2006). A Wnt–Axin2–GSK3β cascade regulates Snail1 activity in breast cancer cells. Nature Cell Biology 8, 1398–1406. 10.1038/ncb1508.

11. Liu, J., Xiao, Q., Xiao, J., Niu, C., Li, Y., Zhang, X., Zhou, Z., Shu, G., and Yin, G. (2022). Wnt/β-catenin signalling: function, biological mechanisms, and therapeutic opportunities. Signal Transduction and Targeted Therapy 7, 3. 10.1038/s41392-021-00762-6.

12. Satoh, S., Daigo, Y., Furukawa, Y., Kato, T., Miwa, N., Nishiwaki, T., Kawasoe, T., Ishiguro, H., Fujita, M., Tokino, T., et al. (2000). AXIN1 mutations in hepatocellular carcinomas, and growth suppression in cancer cells by virus-mediated transfer of AXIN1. Nature Genetics 24, 245–250. 10.1038/73448.

13. Lustig, B., Jerchow, B., Sachs, M., Weiler, S., Pietsch, T., Karsten, U., van de Wetering, M., Clevers, H., Schlag, P.M., Birchmeier, W., and Behrens, J. (2002). Negative Feedback Loop of Wnt Signaling through Upregulation of Conductin/Axin2 in Colorectal and Liver Tumors. Molecular and Cellular Biology 22, 1184–1193. 10.1128/MCB.22.4.1184-1193.2002.

14. Korinek, V., Barker, N., Morin, P.J., van Wichen, D., de Weger, R., Kinzler, K.W., Vogelstein, B., and Clevers, H. (1997). Constitutive Transcriptional Activation by a β-Catenin-Tcf Complex in APC−/− Colon Carcinoma. Science 275, 1784–1787. 10.1126/science.275.5307.1784.

15. Chan, J.M., Clendenning, M., Joseland, S., Georgeson, P., Mahmood, K., Walker, R., Como, J., Joo, J.E., Preston, S., Hutchinson, R.A., et al. (2022). Rare germline variants in the AXIN2 gene in families with colonic polyposis and colorectal cancer. Familial Cancer 21, 399–413. 10.1007/s10689-021-00283-9.

16. Leclerc, J., Beaumont, M., Vibert, R., Pinson, S., Vermaut, C., Flament, C., Lovecchio, T., Delattre, L., Demay, C., Coulet, F., et al. (2023). AXIN2 germline testing in a French cohort validates pathogenic variants as a rare cause of predisposition to colorectal polyposis and cancer. Genes, Chromosomes and Cancer 62, 210–222. 10.1002/gcc.23112.

17. Wu, Z.-Q., Brabletz, T., Fearon, E., Willis, A.L., Hu, C.Y., Li, X.-Y., and Weiss, S.J. (2012). Canonical Wnt suppressor, Axin2, promotes colon carcinoma oncogenic activity. Proceedings of the National Academy of Sciences 109, 11312-11317. 10.1073/pnas.1203015109.

18. Cliffe, A., Hamada, F., and Bienz, M. (2003). A Role of Dishevelled in Relocating Axin to the Plasma Membrane during Wingless Signaling. Current Biology 13, 960–966. 10.1016/S0960-9822(03)00370-1.

19. Ikeda, S., Kishida, S., Yamamoto, H., Murai, H., Koyama, S., and Kikuchi, A. (1998). Axin, a negative regulator of the Wnt signaling pathway, forms a complex with GSK-3β and β-catenin and promotes GSK-3β-dependent phosphorylation of β-catenin. The EMBO Journal 17, 1371–1384-1384. 10.1093/emboj/17.5.1371.

20. Jho, E.-h., Zhang, T., Domon, C., Joo, C.-K., Freund, J.-N., and Costantini, F. (2002). Wnt/β-Catenin/Tcf Signaling Induces the Transcription of Axin2, a Negative Regulator of the Signaling Pathway. Molecular and Cellular Biology 22, 1172–1183. 10.1128/MCB.22.4.1172-1183.2002.

21. Zeng, X., Huang, H., Tamai, K., Zhang, X., Harada, Y., Yokota, C., Almeida, K., Wang, J., Doble, B., Woodgett, J., et al. (2008). Initiation of Wnt signaling: control of Wnt coreceptor Lrp6 phosphorylation/activation via frizzled, dishevelled and axin functions. Development 135, 367–375. 10.1242/dev.013540.

22. Rubinfeld, B., Albert, I., Porfiri, E., Fiol, C., Munemitsu, S., and Polakis, P. (1996). Binding of GSK3β to the APC-β-Catenin Complex and Regulation of Complex Assembly. Science 272, 1023–1026. 10.1126/science.272.5264.1023.

23. Orford, K., Crockett, C., Jensen, J.P., Weissman, A.M., and Byers, S.W. (1997). Serine Phosphorylation-regulated Ubiquitination and Degradation of β-Catenin*. Journal of Biological Chemistry 272, 24735–24738. 10.1074/jbc.272.40.24735.

24. Hart, M., Concordet, J.P., Lassot, I., Albert, I., del los Santos, R., Durand, H., Perret, C., Rubinfeld, B., Margottin, F., Benarous, R., and Polakis, P. (1999). The F-box protein β-TrCP associates with phosphorylated β-catenin and regulates its activity in the cell. Current Biology 9, 207–211. 10.1016/S0960-9822(99)80091-8.

25. Aberle, H., Bauer, A., Stappert, J., Kispert, A., and Kemler, R. (1997). β-catenin is a target for the ubiquitin-proteasome pathway. The EMBO journal 16, 3797–3804. 10.1093/emboj/16.13.3797.

26. Behrens, J., von Kries, J.P., Kühl, M., Bruhn, L., Wedlich, D., Grosschedl, R., and Birchmeier, W. (1996). Functional interaction of β-catenin with the transcription factor LEF-1. Nature 382, 638–642. 10.1038/382638a0.

27. Molenaar, M., van de Wetering, M., Oosterwegel, M., Peterson-Maduro, J., Godsave, S., Korinek, V., Roose, J., Destrée, O., and Clevers, H. (1996). XTcf-3 Transcription Factor Mediates β-catenin-Induced Axis Formation in Xenopus Embryos. Cell 86, 391–399. 10.1016/S0092-8674(00)80112-9.

28. Moshkovsky, A.R., and Kirschner, M.W. (2022). The nonredundant nature of the Axin2 regulatory network in the canonical Wnt signaling pathway. Proceedings of the National Academy of Sciences 119, e2108408119. 10.1073/pnas.2108408119.

29. Zeng, X., Huang, H., Tamai, K., Zhang, X., Harada, Y., Yokota, C., Almeida, K., Wang, J., Doble, B., Woodgett, J., et al. (2008). Initiation of Wnt signaling: control of Wnt coreceptor Lrp6 phosphorylation/activation via frizzled, dishevelled and axin functions. Development 135, 367–375. 10.1242/dev.013540.

30. Mao, J., Wang, J., Liu, B., Pan, W., Farr, G.H., 3rd, Flynn, C., Yuan, H., Takada, S., Kimelman, D., Li, L., and Wu, D. (2001). Low-density lipoprotein receptor-related protein-5 binds to Axin and regulates the canonical Wnt signaling pathway. Mol Cell 7, 801-809. 10.1016/s1097-2765(01)00224-6.

31. Zeng, X., Tamai, K., Doble, B., Li, S., Huang, H., Habas, R., Okamura, H., Woodgett, J., and He, X. (2005). A dual-kinase mechanism for Wnt co-receptor phosphorylation and activation. Nature 438, 873–877. 10.1038/nature04185.

32. Bilic, J., Huang, Y.L., Davidson, G., Zimmermann, T., Cruciat, C.M., Bienz, M., and Niehrs, C. (2007). Wnt induces LRP6 signalosomes and promotes dishevelled-dependent LRP6 phosphorylation. Science 316, 1619–1622. 10.1126/science.1137065.

33. Liu, W., Dong, X., Mai, M., Seelan, R.S., Taniguchi, K., Krishnadath, K.K., Halling, K.C., Cunningham, J.M., Qian, C., Christensen, E., et al. (2000). Mutations in AXIN2 cause colorectal cancer with defective mismatch repair by activating β-catenin/TCF signalling. Nature Genetics 26, 146–147. 10.1038/79859.

34. Bergendal, B., Klar, J., Stecksén-Blicks, C., Norderyd, J., and Dahl, N. (2011). Isolated oligodontia associated with mutations in EDARADD, AXIN2, MSX1, and PAX9 genes. American Journal of Medical Genetics Part A 155, 1616-1622. 10.1002/ajmg.a.34045.

35. Huang, S.-M.A., Mishina, Y.M., Liu, S., Cheung, A., Stegmeier, F., Michaud, G.A., Charlat, O., Wiellette, E., Zhang, Y., Wiessner, S., et al. (2009). Tankyrase inhibition stabilizes axin and antagonizes Wnt signalling. Nature 461, 614–620. 10.1038/nature08356.

36. Qian, L., Mahaffey, J.P., Alcorn, H.L., and Anderson, K.V. (2011). Tissue-specific roles of Axin2 in the inhibition and activation of Wnt signaling in the mouse embryo. Proceedings of the National Academy of Sciences 108, 8692–8697. 10.1073/pnas.1100328108.

37. Morrone, S., Cheng, Z., Moon, R.T., Cong, F., and Xu, W. (2012). Crystal structure of a Tankyrase-Axin complex and its implications for Axin turnover and Tankyrase substrate recruitment. Proceedings of the National Academy of Sciences 109, 1500–1505. doi:10.1073/pnas.1116618109.

38. Landrum, M.J., Lee, J.M., Riley, G.R., Jang, W., Rubinstein, W.S., Church, D.M., and Maglott, D.R. (2014). ClinVar: public archive of relationships among sequence variation and human phenotype. Nucleic Acids Research 42, D980–D985. 10.1093/nar/gkt1113.

39. Yang, Y., Muzny, D.M., Xia, F., Niu, Z., Person, R., Ding, Y., Ward, P., Braxton, A., Wang, M., Buhay, C., et al. (2014). Molecular findings among patients referred for clinical whole-exome sequencing. JAMA 312, 1870–1879. 10.1001/jama.2014.14601.

40. Keehan, L., Jiang, M.M., Li, X., Marom, R., Dai, H., Murdock, D., Liu, P., Hunter, J.V., Heaney, J.D., Robak, L., et al. (2021). A novel de novo intronic variant in ITPR1 causes Gillespie syndrome. Am J Med Genet A 185, 2315–2324. 10.1002/ajmg.a.62232.

41. Chen, S., Francioli, L.C., Goodrich, J.K., Collins, R.L., Kanai, M., Wang, Q., Alföldi, J., Watts, N.A., Vittal, C., Gauthier, L.D., et al. (2022). A genome-wide mutational constraint map quantified from variation in 76,156 human genomes. bioRxiv, 2022.2003.2020.485034-482022.485003.485020.485034. 10.1101/2022.03.20.485034.

42. Goujon, M., McWilliam, H., Li, W., Valentin, F., Squizzato, S., Paern, J., and Lopez, R. (2010). A new bioinformatics analysis tools framework at EMBL–EBI. Nucleic Acids Research 38, W695–W699. 10.1093/nar/gkq313.

43. Sievers, F., Wilm, A., Dineen, D., Gibson, T.J., Karplus, K., Li, W., Lopez, R., McWilliam, H., Remmert, M., Söding, J., et al. (2011). Fast, scalable generation of high-quality protein multiple sequence alignments using Clustal Omega. Molecular Systems Biology 7, 539. 10.1038/msb.2011.75.

44. Söding, J. (2005). Protein homology detection by HMM–HMM comparison. Bioinformatics 21, 951–960. 10.1093/bioinformatics/bti125.

45. UniProt, C. (2023). UniProt: the Universal Protein Knowledgebase in 2023. Nucleic Acids Res 51, D523–D531. 10.1093/nar/gkac1052.

46. Emsley, P., Lohkamp, B., Scott, W.G., and Cowtan, K. (2010). Features and development of Coot. Acta Crystallogr D Biol Crystallogr 66, 486–501. 10.1107/S0907444910007493.

47. Hodgkins, A., Farne, A., Perera, S., Grego, T., Parry-Smith, D.J., Skarnes, W.C., and Iyer, V. (2015). WGE: a CRISPR database for genome engineering. Bioinformatics 31, 3078–3080. 10.1093/bioinformatics/btv308.

48. Lanza, D.G., Mao, J., Lorenzo, I., Liao, L., Seavitt, J.R., Ljungberg, M.C., Simpson, E.M., DeMayo, F.J., and Heaney, J.D. (2024). An oocyte-specific Cas9-expressing mouse for germline CRISPR/Cas9-mediated genome editing. Genesis 62, e23589. 10.1002/dvg.23589.

49. Chow, R.D., Chen, J.S., Shen, J., and Chen, S. (2021). A web tool for the design of prime-editing guide RNAs. Nat Biomed Eng 5, 190–194. 10.1038/s41551-020-00622-8.

50. Hai, L., Szwarc, M.M., Lanza, D.G., Heaney, J.D., and Lydon, J.P. (2019). Using CRISPR/Cas9 engineering to generate a mouse with a conditional knockout allele for the promyelocytic leukemia zinc finger transcription factor. Genesis 57, e23281. 10.1002/dvg.23281.

51. Nakao, K., Nakagata, N., and Katsuki, M. (1997). Simple and Efficient Vitrification Procedure for Cryopreservation of Mouse Embryos. Experimental Animals 46, 231–234. 10.1538/expanim.46.231.

52. Nakagata, N., and Takeo, T. (2019). Basic mouse reproductive techniques developed and modified at the Center for Animal Resources and Development (CARD), Kumamoto University. Experimental Animals 68, 391–395. 10.1538/expanim.19-0070.

53. Dickinson, M.E., Flenniken, A.M., Ji, X., Teboul, L., Wong, M.D., White, J.K., Meehan, T.F., Weninger, W.J., Westerberg, H., Adissu, H., et al. (2016). High-throughput discovery of novel developmental phenotypes. Nature 537, 508–514. 10.1038/nature19356.

54. Takeo, T., Hoshii, T., Kondo, Y., Toyodome, H., Arima, H., Yamamura, K.-i., Irie, T., and Nakagata, N. (2008). Methyl-Beta-Cyclodextrin Improves Fertilizing Ability of C57BL/6 Mouse Sperm after Freezing and Thawing by Facilitating Cholesterol Efflux from the Cells1. Biology of Reproduction 78, 546–551. 10.1095/biolreprod.107.065359.

55. Takeo, T., and Nakagata, N. (2011). Reduced Glutathione Enhances Fertility of Frozen/Thawed C57BL/6 Mouse Sperm after Exposure to Methyl-Beta-Cyclodextrin1. Biology of Reproduction 85, 1066–1072. 10.1095/biolreprod.111.092536.

56. Chung, H.L., Mao, X., Wang, H., Park, Y.J., Marcogliese, P.C., Rosenfeld, J.A., Burrage, L.C., Liu, P., Murdock, D.R., Yamamoto, S., et al. (2020). De Novo Variants in CDK19 Are Associated with a Syndrome Involving Intellectual Disability and Epileptic Encephalopathy. Am J Hum Genet 106, 717–725. 10.1016/j.ajhg.2020.04.001.

57. Bischof, J., Bjorklund, M., Furger, E., Schertel, C., Taipale, J., and Basler, K. (2013). A versatile platform for creating a comprehensive UAS-ORFeome library in Drosophila. Development 140, 2434–2442. 10.1242/dev.088757.

58. Venken, K.J., He, Y., Hoskins, R.A., and Bellen, H.J. (2006). P[acman]: a BAC transgenic platform for targeted insertion of large DNA fragments in D. melanogaster. Science 314, 1747–1751. 10.1126/science.1134426.

59. Kanca, O., Zirin, J., Hu, Y., Tepe, B., Dutta, D., Lin, W.W., Ma, L., Ge, M., Zuo, Z., Liu, L.P., et al. (2022). An expanded toolkit for Drosophila gene tagging using synthesized homology donor constructs for CRISPR-mediated homologous recombination. Elife 11. 10.7554/eLife.76077.

60. Marcogliese, P.C., Dutta, D., Ray, S.S., Dang, N.D.P., Zuo, Z., Wang, Y., Lu, D., Fazal, F., Ravenscroft, T.A., Chung, H., et al. (2022). Loss of IRF2BPL impairs neuronal maintenance through excess Wnt signaling. Sci Adv 8, eabl5613. 10.1126/sciadv.abl5613.

61. Nolo, R., Abbott, L.A., and Bellen, H.J. (2000). Senseless, a Zn finger transcription factor, is necessary and sufficient for sensory organ development in Drosophila. Cell 102, 349–362. 10.1016/s0092-8674(00)00040-4.

62. Rentzsch, P., Schubach, M., Shendure, J., and Kircher, M. (2021). CADD-Splice-improving genome-wide variant effect prediction using deep learning-derived splice scores. Genome Med 13, 31. 10.1186/s13073-021-00835-9.

63. Ioannidis, N.M., Rothstein, J.H., Pejaver, V., Middha, S., McDonnell, S.K., Baheti, S., Musolf, A., Li, Q., Holzinger, E., Karyadi, D., et al. (2016). REVEL: An Ensemble Method for Predicting the Pathogenicity of Rare Missense Variants. Am J Hum Genet 99, 877–885. 10.1016/j.ajhg.2016.08.016.

64. Cheng, J., Novati, G., Pan, J., Bycroft, C., Zemgulyte, A., Applebaum, T., Pritzel, A., Wong, L.H., Zielinski, M., Sargeant, T., et al. (2023). Accurate proteome-wide missense variant effect prediction with AlphaMissense. Science 381, eadg7492. 10.1126/science.adg7492.

65. Morrone, S., Cheng, Z., Moon, R.T., Cong, F., and Xu, W. (2012). Crystal structure of a Tankyrase-Axin complex and its implications for Axin turnover and Tankyrase substrate recruitment. Proc Natl Acad Sci U S A 109, 1500–1505. 10.1073/pnas.1116618109.

66. Sondka, Z., Dhir, N.B., Carvalho-Silva, D., Jupe, S., Madhumita, McLaren, K., Starkey, M., Ward, S., Wilding, J., Ahmed, M., et al. (2024). COSMIC: a curated database of somatic variants and clinical data for cancer. Nucleic Acids Res 52, D1210–D1217. 10.1093/nar/gkad986.

67. Evans, R., O’Neill, M., Pritzel, A., Antropova, N., Senior, A., Green, T., Žídek, A., Bates, R., Blackwell, S., Yim, J., et al. (2022). Protein complex prediction with AlphaFold-Multimer. bioRxiv, 2021.2010.2004.463034. 10.1101/2021.10.04.463034.

68. Abramson, J., Adler, J., Dunger, J., Evans, R., Green, T., Pritzel, A., Ronneberger, O., Willmore, L., Ballard, A.J., Bambrick, J., et al. (2024). Accurate structure prediction of biomolecular interactions with AlphaFold 3. Nature 630, 493–500. 10.1038/s41586-024-07487-w.

69. Guettler, S., LaRose, J., Petsalaki, E., Gish, G., Scotter, A., Pawson, T., Rottapel, R., and Sicheri, F. (2011). Structural Basis and Sequence Rules for Substrate Recognition by Tankyrase Explain the Basis for Cherubism Disease. Cell 147, 1340–1354. 10.1016/j.cell.2011.10.046.

70. Teboul, L., Murray, S.A., and Nolan, P.M. (2017). Phenotyping first-generation genome editing mutants: a new standard? Mamm Genome 28, 377–382. 10.1007/s00335-017-9711-x.

71. Liu, Y., Li, X., He, S., Huang, S., Li, C., Chen, Y., Liu, Z., Huang, X., and Wang, X. (2020). Efficient generation of mouse models with the prime editing system. Cell Discov 6, 27. 10.1038/s41421-020-0165-z.

72. Adikusuma, F., Lushington, C., Arudkumar, J., Godahewa, G.I., Chey, Y.C.J., Gierus, L., Piltz, S., Geiger, A., Jain, Y., Reti, D., et al. (2021). Optimized nickase- and nuclease-based prime editing in human and mouse cells. Nucleic Acids Res 49, 10785–10795. 10.1093/nar/gkab792.

73. Park, S.J., Jeong, T.Y., Shin, S.K., Yoon, D.E., Lim, S.Y., Kim, S.P., Choi, J., Lee, H., Hong, J.I., Ahn, J., et al. (2021). Targeted mutagenesis in mouse cells and embryos using an enhanced prime editor. Genome Biol 22, 170. 10.1186/s13059-021-02389-w.

74. Gao, P., Lyu, Q., Ghanam, A.R., Lazzarotto, C.R., Newby, G.A., Zhang, W., Choi, M., Slivano, O.J., Holden, K., Walker, J.A., 2nd, et al. (2021). Prime editing in mice reveals the essentiality of a single base in driving tissue-specific gene expression. Genome Biol 22, 83. 10.1186/s13059-021-02304-3.

75. Her, Y., Pascual, D.M., Goldstone-Joubert, Z., and Marcogliese, P.C. (2024). Variant functional assessment in Drosophila by overexpression: what can we learn? Genome 67, 158–167. 10.1139/gen-2023-0135.

76. Swarup, S., and Verheyen, E.M. (2012). Wnt/Wingless signaling in Drosophila. Cold Spring Harb Perspect Biol 4. 10.1101/cshperspect.a007930.

77. Nagarkar-Jaiswal, S., Lee, P.-T., Campbell, M.E., Chen, K., Anguiano-Zarate, S., Cantu Gutierrez, M., Busby, T., Lin, W.-W., He, Y., Schulze, K.L., et al. (2015). A library of MiMICs allows tagging of genes and reversible, spatial and temporal knockdown of proteins in Drosophila. eLife 4, e05338. 10.7554/eLife.05338.

78. Roht, L., Hyldebrandt, H.K., Stormorken, A.T., Nordgarden, H., Sijmons, R.H., Bos, D.K., Riegert-Johnson, D., Mantia-Macklin, S., Ilves, P., Muru, K., et al. (2023). AXIN2-related oligodontia-colorectal cancer syndrome with cleft palate as a possible new feature. Molecular Genetics & Genomic Medicine 11, e2157. 10.1002/mgg3.2157.

79. Macklin-Mantia, S.K., Hines, S.L., Chaichana, K.L., Donaldson, A.M., Ko, S.L., Zhai, Q., Samadder, N.J., and Riegert-Johnson, D.L. (2020). Case report expanding the germline AXIN2-related phenotype to include olfactory neuroblastoma and gastric adenoma. BMC Medical Genetics 21, 161. 10.1186/s12881-020-01103-0.

80. Yu, H.-M.I., Jerchow, B., Sheu, T.-J., Liu, B., Costantini, F., Puzas, J.E., Birchmeier, W., and Hsu, W. (2005). The role of Axin2 in calvarial morphogenesis and craniosynostosis. Development 132, 1995–2005. 10.1242/dev.01786.

81. Yan, Y., Tang, D., Chen, M., Huang, J., Xie, R., Jonason, J.H., Tan, X., Hou, W., Reynolds, D., Hsu, W., et al. (2009). Axin2 controls bone remodeling through the β-catenin–BMP signaling pathway in adult mice. Journal of Cell Science 122, 3566–3578. 10.1242/jcs.051904.

82. de Roo, J.J.D., Breukel, C., Chhatta, A.R., Linssen, M.M., Vloemans, S.A., Salvatori, D., Mikkers, H.M.M., Verbeek, S.J., and Staal, F.J.T. (2017). Axin2-mTurquoise2: A novel reporter mouse model for the detection of canonical Wnt signalling. Genesis 55, e23068. 10.1002/dvg.23068.

83. Mazzoni, S.M., Petty, E.M., Stoffel, E.M., and Fearon, E.R. (2015). An AXIN2 Mutant Allele Associated With Predisposition to Colorectal Neoplasia Has Context-Dependent Effects on AXIN2 Protein Function. Neoplasia 17, 463–472. 10.1016/j.neo.2015.04.006.

84. Janeckova, E., Feng, J., Guo, T., Han, X., Ghobadi, A., Araujo-Villalba, A., Rahman, M.S., Ziaei, H., Ho, T.V., Pareek, S., et al. (2023). Canonical Wnt signaling regulates soft palate development by mediating ciliary homeostasis. Development 150. 10.1242/dev.201189.

85. Iwata, J., Suzuki, A., Yokota, T., Ho, T.V., Pelikan, R., Urata, M., Sanchez-Lara, P.A., and Chai, Y. (2014). TGFbeta regulates epithelial-mesenchymal interactions through WNT signaling activity to control muscle development in the soft palate. Development 141, 909–917. 10.1242/dev.103093.

86. Maupin, K.A., Droscha, C.J., and Williams, B.O. (2013). A Comprehensive Overview of Skeletal Phenotypes Associated with Alterations in Wnt/beta-catenin Signaling in Humans and Mice. Bone Res 1, 27–71. 10.4248/BR201301004.

87. Micchelli, C.A., Rulifson, E.J., and Blair, S.S. (1997). The function and regulation of cut expression on the wing margin of Drosophila: Notch, Wingless and a dominant negative role for Delta and Serrate. Development 124, 1485–1495. 10.1242/dev.124.8.1485.

88. Legent, K., and Treisman, J.E. (2008). Wingless signaling in Drosophila eye development. Methods Mol Biol 469, 141–161. 10.1007/978-1-60327-469-2_12.

