## Supplemental Material for "Uncovering Phenotypic Expansion in AXIN2-Related Disorders through Precision Animal Modeling"

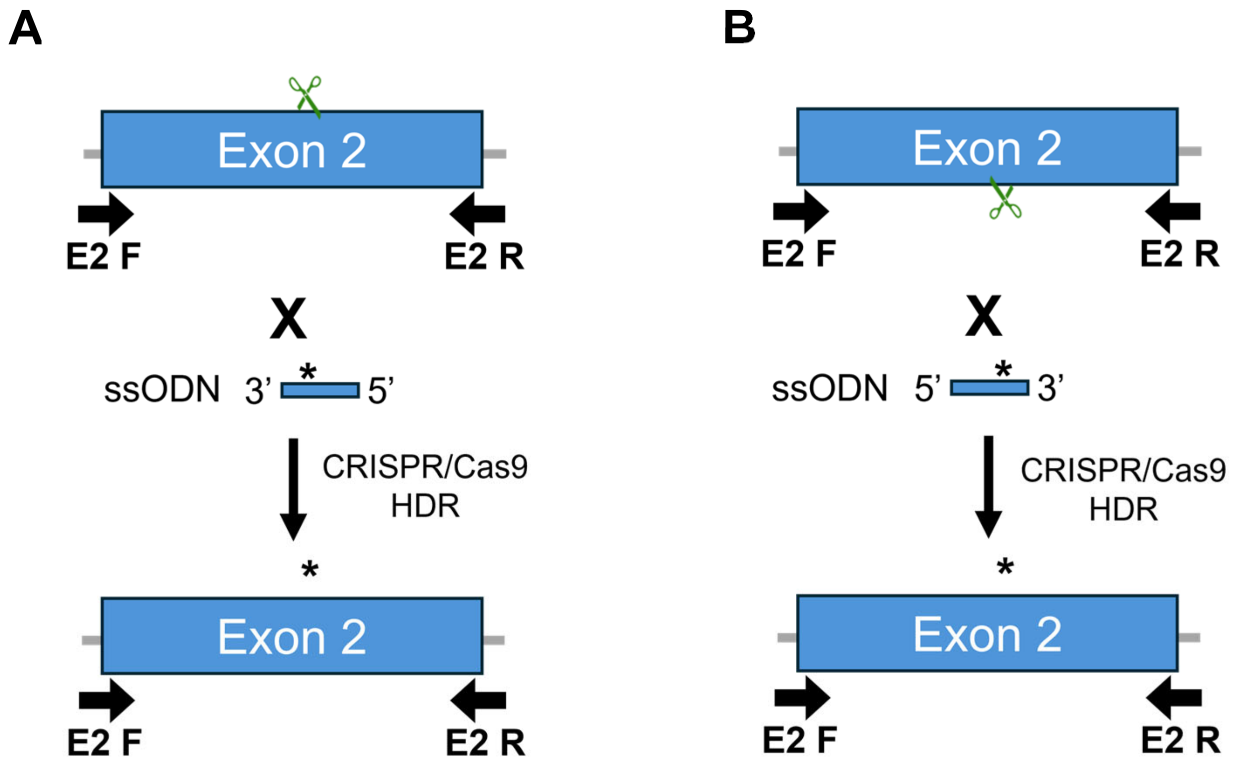
A
Exon 2
E2 F
E2 R
X
ssODN 3' * 5'
CRISPR/Cas9
HDR
*
Exon 2
E2 F
E2 R
B
Exon 2
E2 F
E2 R
X
ssODN 5' * 3'
CRISPR/Cas9
HDR
*
Exon 2
E2 F
E2 R

Figure S1

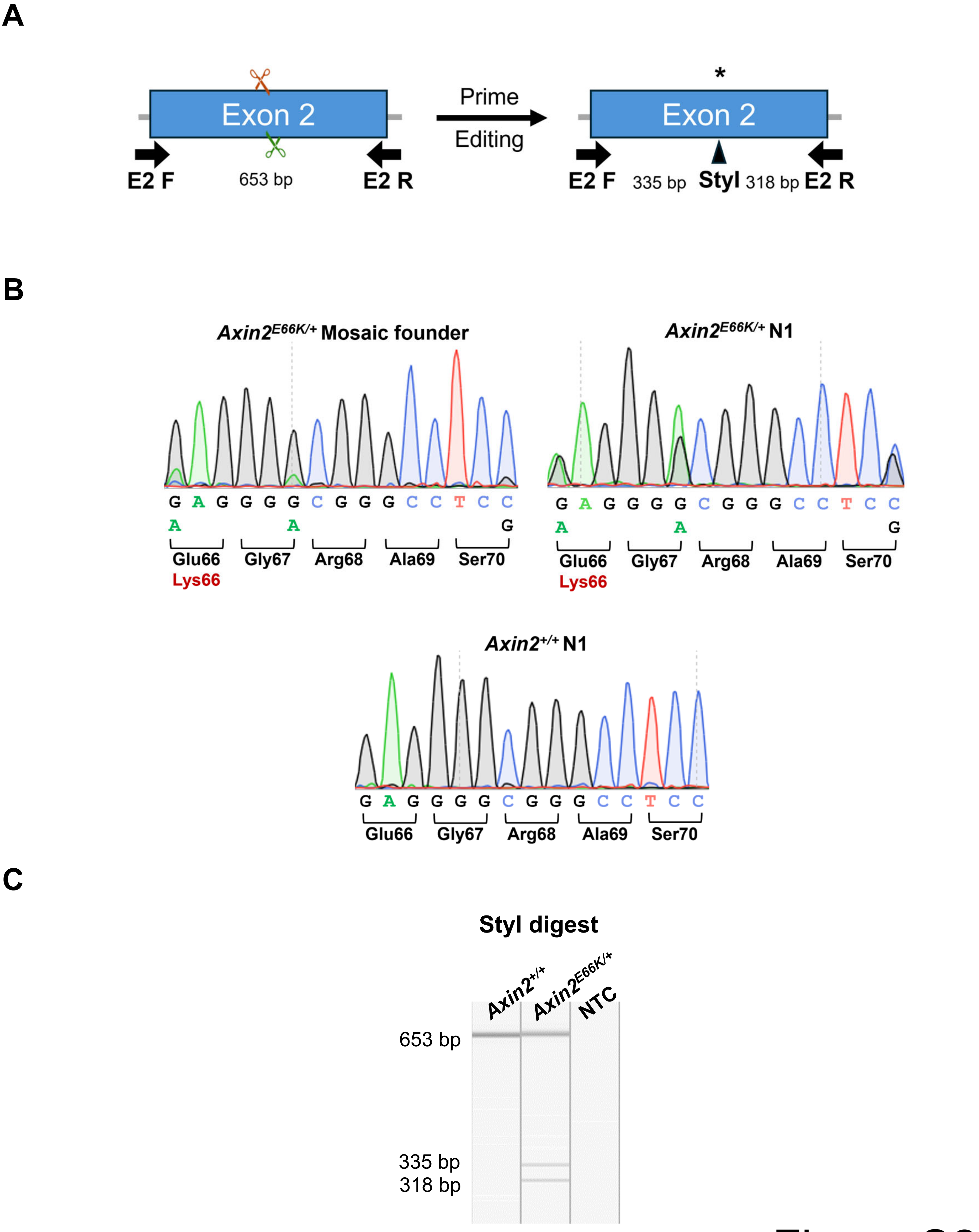
A
Exon 2
E2 F
653 bp
E2 R
Prime
Editing
*
Exon 2
E2 F
335 bp
StyI
318 bp
E2 R
B
Axin2E66K/+ Mosaic founder
G A G G G G C G G G C C T C C
A A G
Glu66 Gly67 Arg68 Ala69 Ser70
Lys66
Axin2E66K/+ N1
G A G G G G C G G G C C T C C
A A G
Glu66 Gly67 Arg68 Ala69 Ser70
Lys66
Axin2+/+ N1
G A G G G G C G G G C C T C C
Glu66 Gly67 Arg68 Ala69 Ser70
C
StyI digest
Axin2+/+
Axin2E66K/+
NTC
653 bp
335 bp
318 bp

Figure S2

**A**

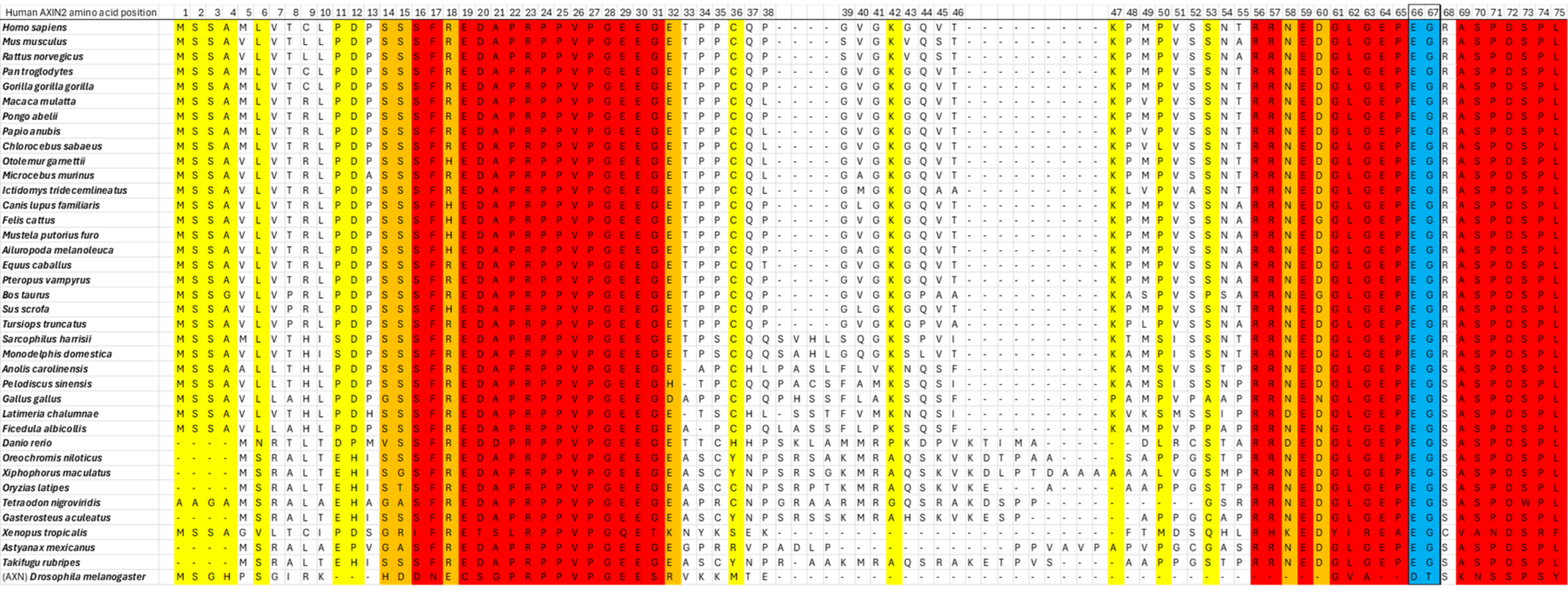

**B**

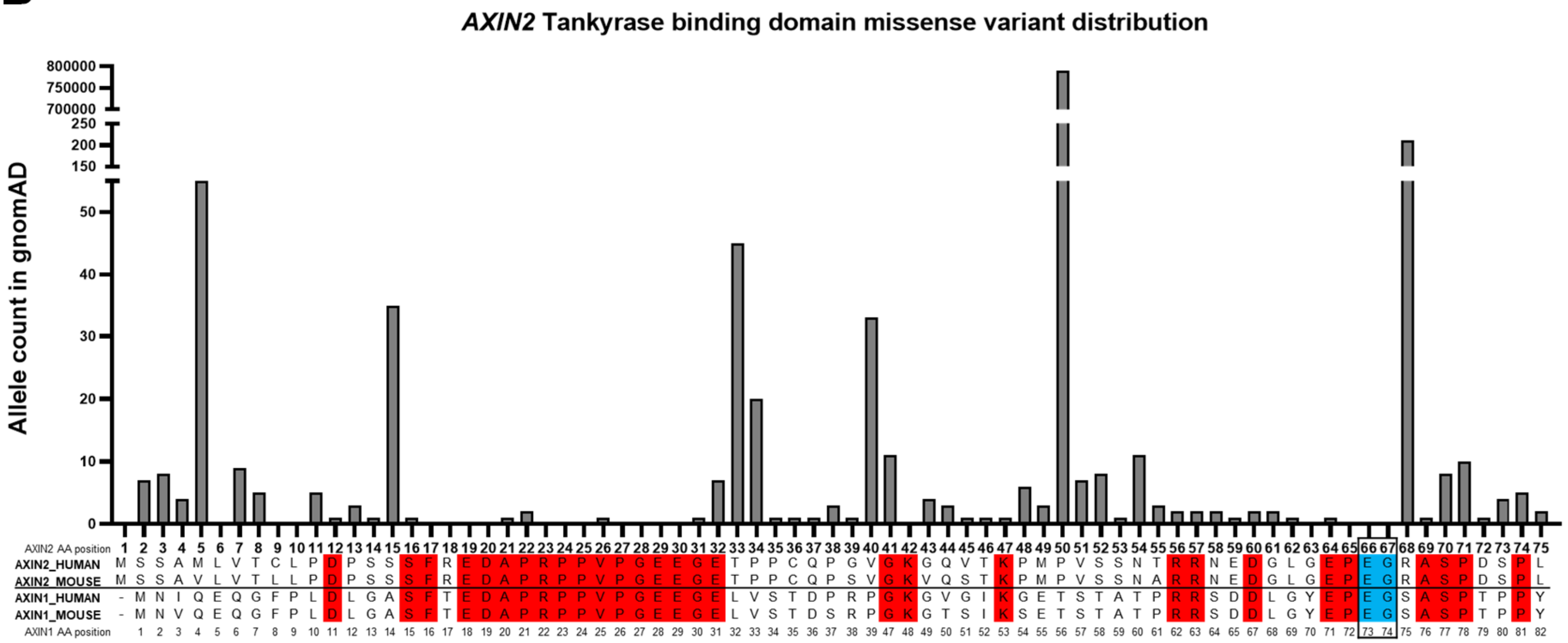

*AXIN2* Tankyrase binding domain missense variant distribution
Allele count in gnomAD
AXIN2 AA position
AXIN2_HUMAN
AXIN2_MOUSE
AXIN1_HUMAN
AXIN1_MOUSE
AXIN1 AA position


Figure S3

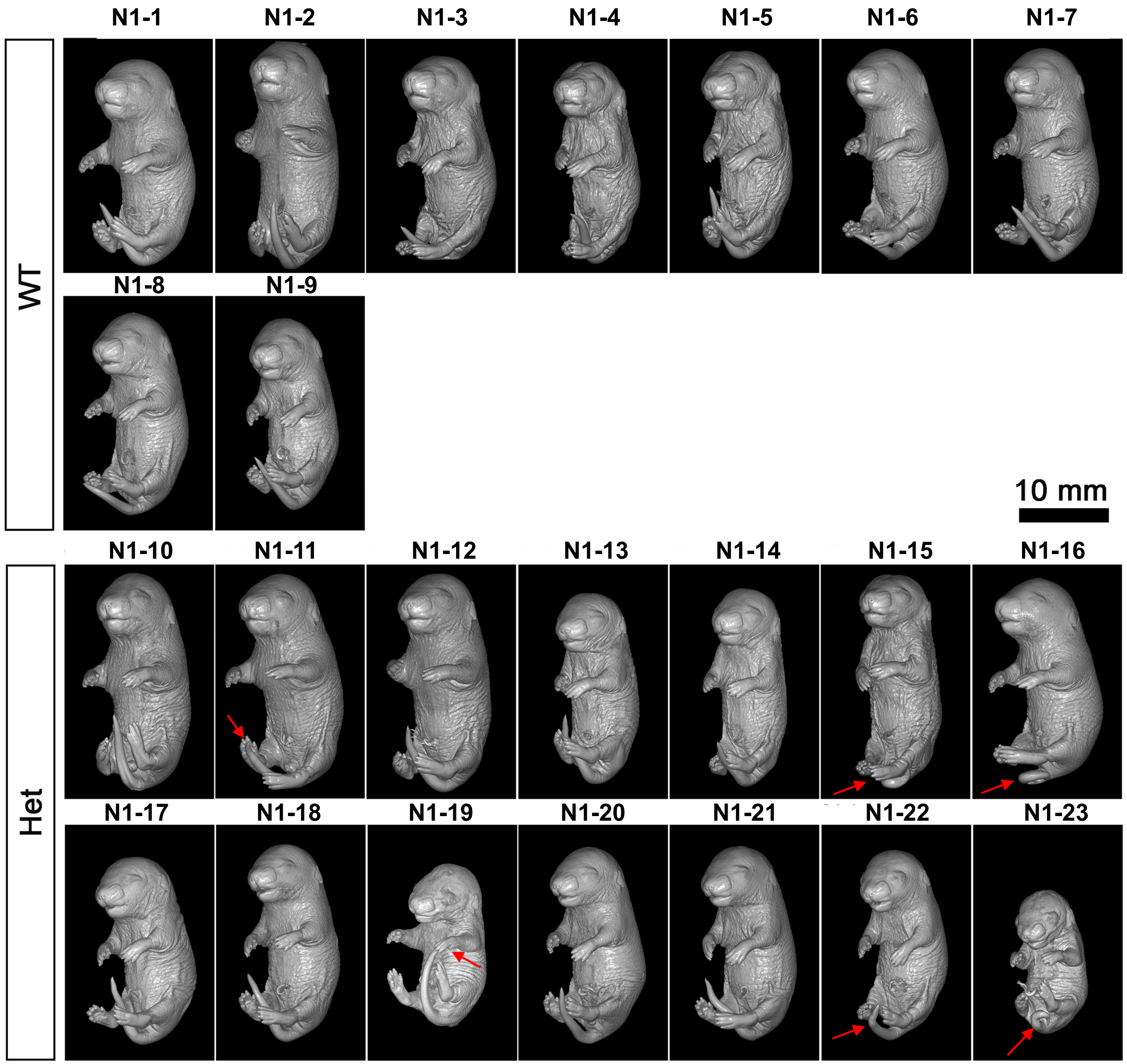

N1-1
N1-2
N1-3
N1-4
N1-5
N1-6
N1-7
WT
N1-8
N1-9
10 mm
N1-10
N1-11
N1-12
N1-13
N1-14
N1-15
N1-16
Het
N1-17
N1-18
N1-19
N1-20
N1-21
N1-22
N1-23


Figure S4

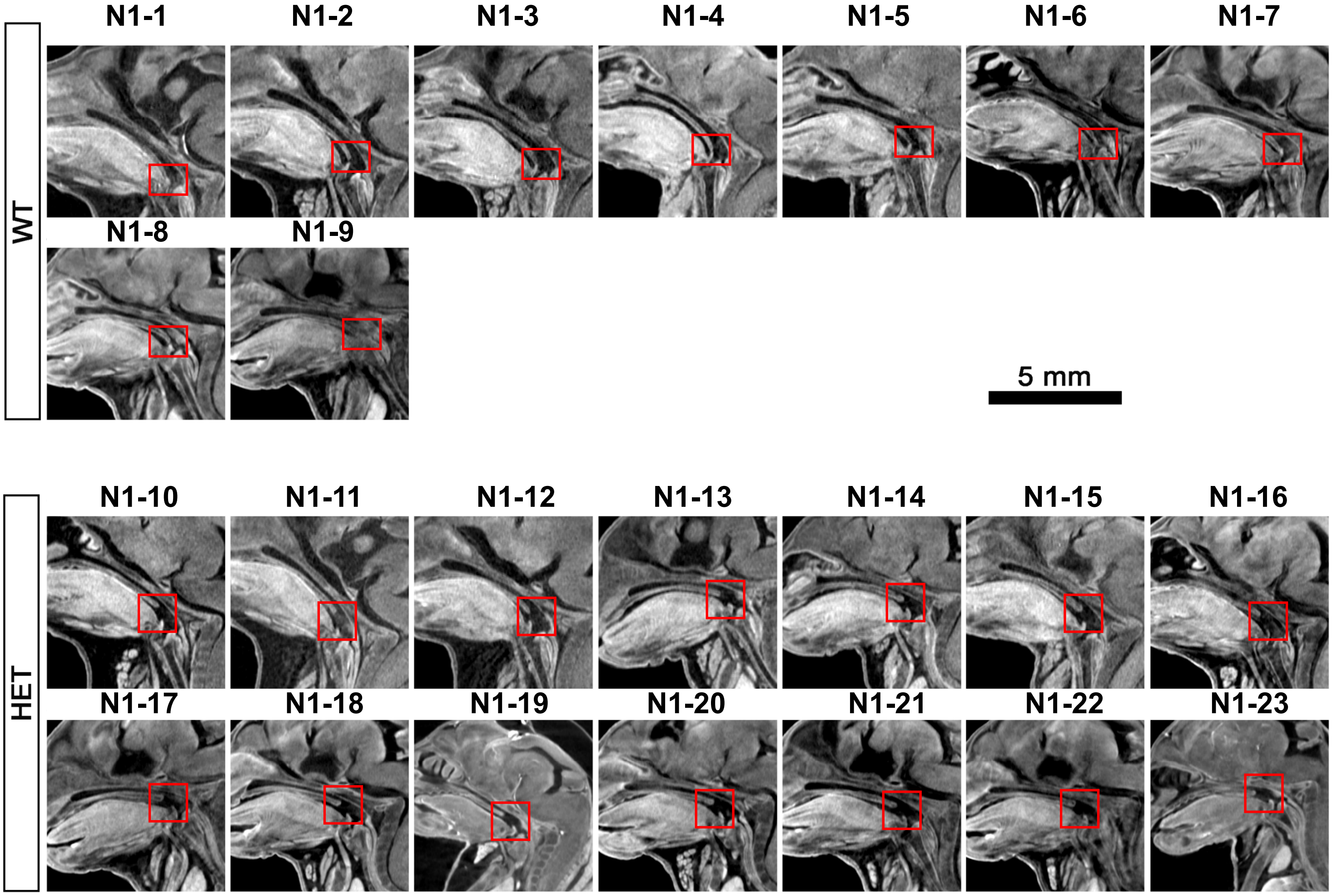
WT
N1-1
N1-2
N1-3
N1-4
N1-5
N1-6
N1-7
N1-8
N1-9
5 mm
HET
N1-10
N1-11
N1-12
N1-13
N1-14
N1-15
N1-16
N1-17
N1-18
N1-19
N1-20
N1-21
N1-22
N1-23

Figure S5

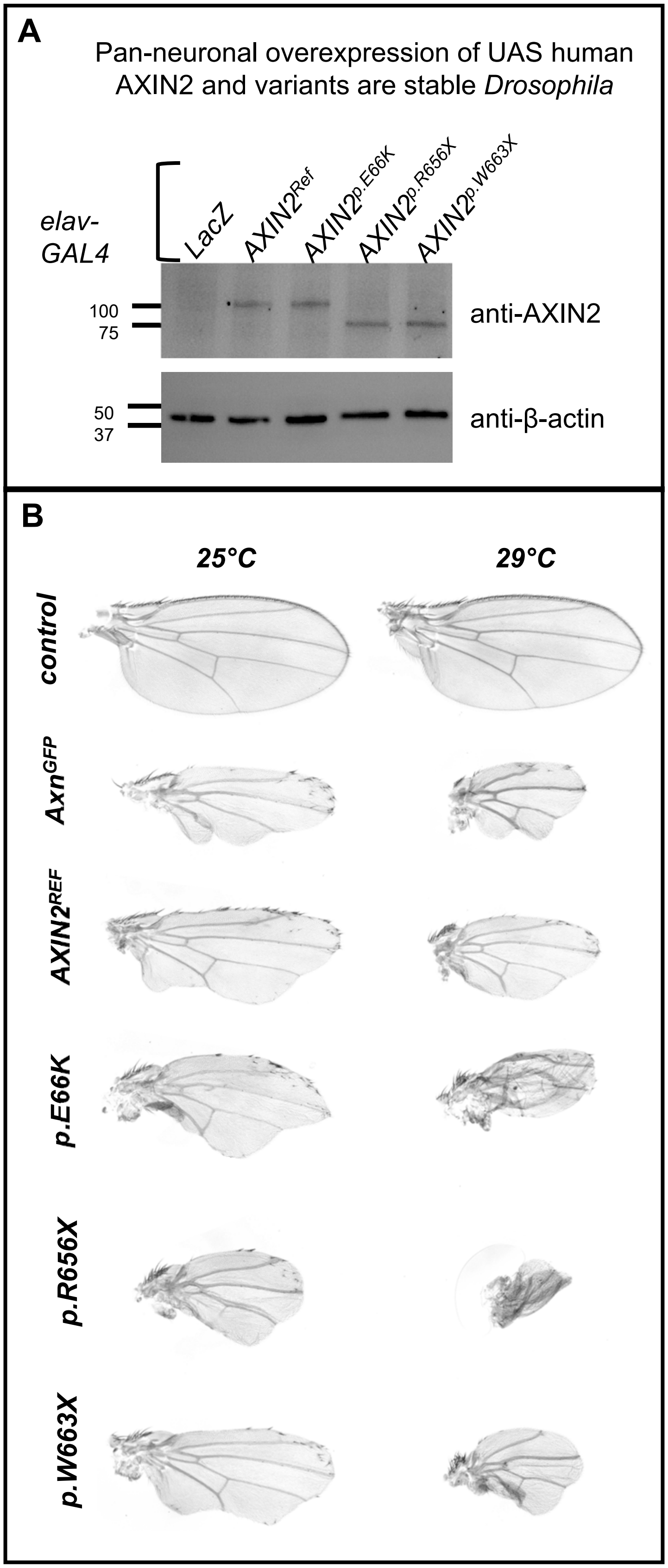
A
Pan-neuronal overexpression of UAS human AXIN2 and variants are stable Drosophila
elav-GAL4
LacZ
AXIN2Ref
AXIN2p.E66K
AXIN2p.R656X
AXIN2p.W663X
100
75
anti-AXIN2
50
37
anti-β-actin
B
25°C
29°C
control
AxnGFP
AXIN2REF
p.E66K
p.R656X
p.W663X

Figure S6

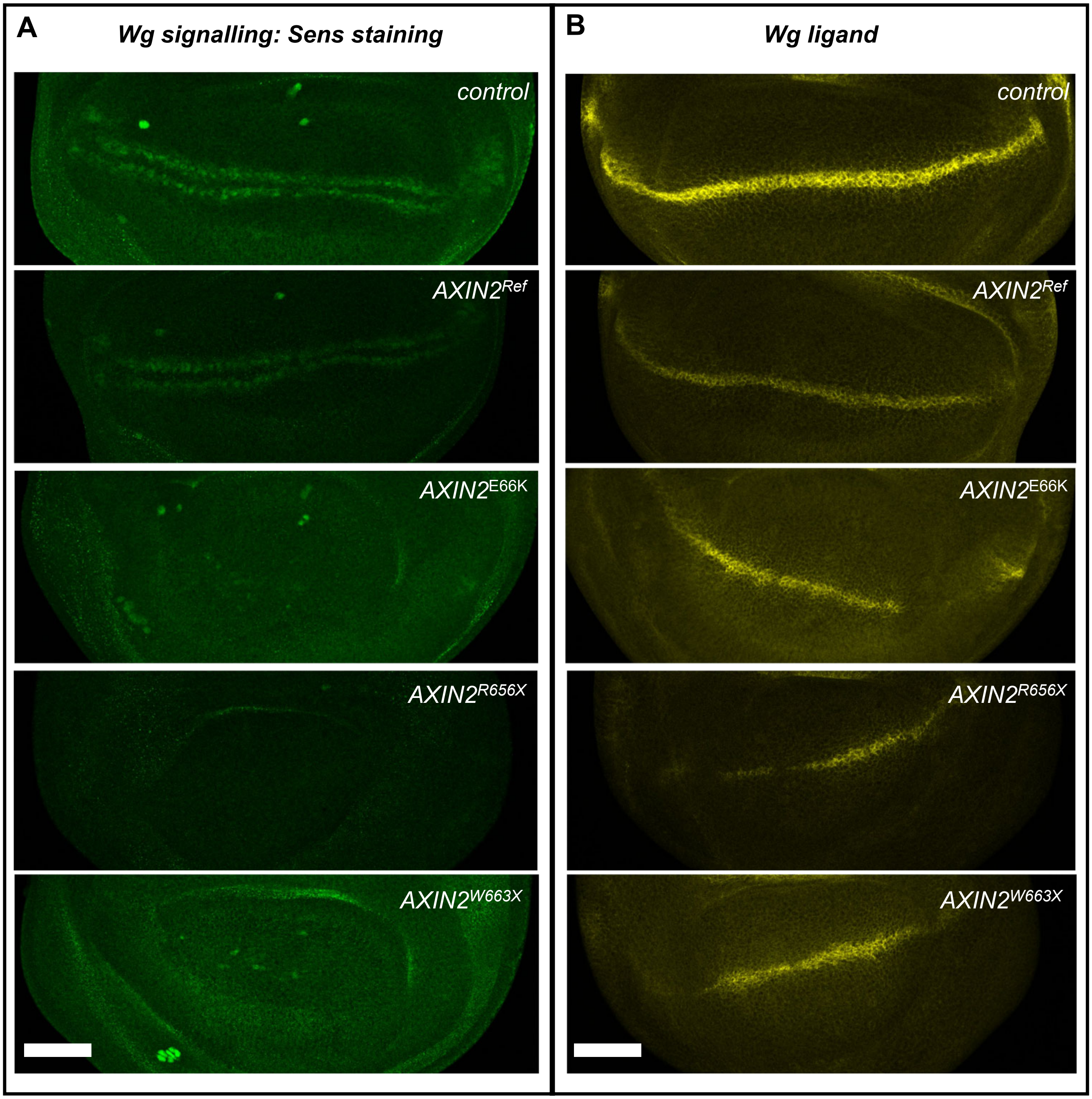
A
Wg signalling: Sens staining
control
AXIN2Ref
AXIN2E66K
AXIN2R656X
AXIN2W663X
B
Wg ligand
control
AXIN2Ref
AXIN2E66K
AXIN2R656X
AXIN2W663X

Figure S7

**Table S1.** Guides for *Axin2* founder screen by traditional CRISPR HDR.

| Targeting Scheme | Component | Chromosome Location* | Sequence (5' to 3')† | PAM | Strand |
|---|---|---|---|---|---|
| Sense | gRNA | 11:108923466-108923488 | ATGGACTGGGGGAGCCCGAG | GGG | + |
| | ssDNA | 11:108923447-108923579 | AAGTCCGGAAGAGGTATGCAC<br>CATCCTGGTCACCCAACAAGG<br>AGTGTAAAGACTTGGTCCACC<br>TGGTCAAAGGGGAATCGGGGG<br>AGGCCCGGCC**CTT**GGGCTCCC<br>CCAGTCCATCTTCATTCCGCC<br>TAGCATT | | - |
| Antisense | gRNA | 11:108923480-108923502 | TCGGGGGAGGCCCGCCCCTC | GGG | - |
| | ssDNA | 11:108923391-108923524 | CGTGTCAGCCTAGTGTGGGCA<br>AGGTCCAGTCCACCAAACCTA<br>TGCCCGTTTCCTCTAATGCTA<br>GGCGGAATGAAGATGGACTGG<br>GGGAGCCC**AAG**GGCCGGGCCT<br>CCCCCGATTCCCCTTTGACCA<br>GGTGGACC | | + |

*Genome build GRCm38
†bold bases denote GAG to AAG p.E66K mutation, underline bases denote GGG to GGC p.G67G synonymous mutation

**Table S2.** Guides for *Axin2* mouse model production by prime editing.

| Allele | Guide ID | Chromosome Location* | Target Sequence (5' to 3') | PAM | Strand |
|---|---|---|---|---|---|
| Prime editing for p.E66K | pegRNA | 11:108923466-108923488 | ATGGACTGGGGGAGCCCGAG | GGG | + |
| | Nicking gRNA | 11:108923523-108923545 | CAACAAGGAGTGTAAAGACT | TGG | - |

*Genome build GRCm38

**Table S3.** Prime editing pegRNA sequences.

| Component | Length | Sequence (5' to 3') |
|---|---|---|
| Target | 20 nt | ATGGACTGGGGGAGCCCGAG |
| RT template | 14 nt | GAGGCCCGTCCCTT |
| PBS | 10 nt | GGGCTCCCCC |
| Sense 3’ extension | 24 nt | GAGGCCCGTCCCTTGGGCTCCCCC |
| Full length pegRNA | 120 nt | ATGGACTGGG GGAGCCCGAG GTTTTAGAGC TAGAAATAGC<br>AAGTTAAAAT AAGGCTAGTC CGTTATCAAC TTGAAAAAGT<br>GGCACCGAGT CGGTGCGAGG CCCGTCCCTT GGGCTCCCCC |

**Table S4.** Primers used for site-directed mutagenesis of AXIN2 for fly modeling.

| Primer | Sequence (5' to 3') |
|---|---|
| AXIN2 p.E66K F | GGGGGAGCCGAAGGGGCGGGC |
| AXIN2 p.E66K R | AACCCATCTTCGTTCCGCCTGGTGTTGGAAGAGAC |
| AXIN2 p.R656X F | TCCAGGCGAATGAGCCAGCCG |
| AXIN2 p.R656X R | GACGAGCGGGCAGACTCC |
| AXIN2 p.W663X F | CACCATCTGTAGGGGGGCAACAGC |
| AXIN2 p.W663X R | CCGGCTGGCTCGTTCGCC |

**Table S5.** Results of ICE analysis of Sanger sequencing reads from p.E66K founder embryos.*

| Embryo ID | Wild-type (%) | p.E66K Knock-In (%) | Indel (%) | Targeting scheme |
|---|---|---|---|---|
| F0-1 | 0 | 0 | 100 | sense |
| F0-2 | 0 | 0 | 100 | sense |
| **F0-3** | **100** | **0** | **0** | **sense** |
| F0-4 | 0 | 0 | 100 | sense |
| F0-5 | 0 | 0 | 100 | sense |
| F0-6 | 0 | 28 | 72 | sense |
| F0-7 | 82 | 18 | 0 | sense |
| F0-8 | 0 | 0 | 100 | sense |
| F0-9 | 0 | 0 | 100 | sense |
| **F0-10** | **0** | **96** | **0** | **sense** |
| F0-11 | 0 | 0 | 100 | sense |
| F0-12 | 0 | 50 | 50 | sense |
| F0-13 | 0 | 42 | 58 | sense |
| F0-14 | 0 | 12 | 88 | sense |
| F0-15 | 0 | 37 | 63 | sense |
| F0-16 | 0 | 0 | 97 | sense |
| F0-17 | 0 | 0 | 100 | sense |
| F0-18 | 0 | 0 | 100 | sense |
| F0-19 | 0 | 7 | 93 | sense |
| F0-20 | 0 | 43 | 57 | sense |
| F0-21 | 0 | 0 | 100 | antisense |
| F0-22 | 0 | 0 | 100 | antisense |
| F0-23 | 0 | 0 | 100 | antisense |
| F0-24 | 0 | 20 | 80 | antisense |
| F0-25 | 0 | 0 | 100 | antisense |
| F0-26 | 67 | 33 | 0 | antisense |
| **F0-27** | **47** | **53** | **0** | **antisense** |
| F0-28 | 100 | 0 | 0 | antisense |
| F0-29 | 50 | 0 | 50 | antisense |
| F0-30 | 100 | 0 | 0 | antisense |
| F0-31 | 50 | 0 | 50 | antisense |
| F0-32 | 100 | 0 | 0 | antisense |
| F0-33 | 50 | 0 | 50 | antisense |
| F0-34 | 66 | 34 | 0 | antisense |
| F0-35 | 100 | 0 | 0 | antisense |
| F0-36 | 50 | 0 | 50 | antisense |
| F0-37 | 82 | 18 | 0 | antisense |

Wild-type, p.E66K knock-in, and indel allele contribution in founder screen embryos. Data from embryos F0-3, F0-10, and F0-27 are included in Figure 3B-D.

**Members of the Undiagnosed Diseases Network (Version: 10.24.2024)**

| Full Name | Affiliation | Email |
|---|---|---|
| Alyssa A. Tran | BCM Clinical | |
| Arjun Tarakad | BCM Clinical | |
| Ashok Balasubramanyam | BCM Clinical | |
| Brendan H. Lee | BCM Clinical | |
| Carlos A. Bacino | BCM Clinical | |
| Daryl A. Scott | BCM Clinical | |
| Elaine Seto | BCM Clinical | |
| Gary D. Clark | BCM Clinical | |
| Hongzheng Dai | BCM Clinical | |
| Hsiao-Tuan Chao | BCM Clinical | |
| Ivan Chinn | BCM Clinical | |
| James P. Orengo | BCM Clinical | |
| Jennifer E. Posey | BCM Clinical | |
| Jill A. Rosenfeld | BCM Clinical | |
| Kim Worley | BCM Clinical | |
| Lindsay C. Burrage | BCM Clinical | |
| Lisa T. Emrick | BCM Clinical | |
| Lorraine Potocki | BCM Clinical | |
| Monika Weisz Hubshman | BCM Clinical | |
| Richard A. Lewis | BCM Clinical | |
| Ronit Marom | BCM Clinical | |
| Seema R. Lalani | BCM Clinical | |
| Shamika Ketkar | BCM Clinical | |
| Tiphanie P. Vogel | BCM Clinical | |
| William J. Craigen | BCM Clinical | |
| Lauren Blieden | BCM Clinical | |
| Jared Sninsky | BCM Clinical | |
| Hugo J. Bellen | BCM MOSC | |
| Michael F. Wangler | BCM MOSC | |
| Oguz Kanca | BCM MOSC | |
| Shinya Yamamoto | BCM MOSC | |
| Christine M. Eng | BCM Sequencing | |
| Patricia A. Ward | BCM Sequencing | |
| Pengfei Liu | BCM Sequencing | |

| | | |
|---|---|---|
| Adeline Vanderver | CHOP | |
| Cara Skraban | CHOP | |
| Edward Behrens | CHOP | |
| Gonench Kilich | CHOP | |
| Kathleen Sullivan | CHOP | |
| Kelly Hassey | CHOP | |
| Ramakrishnan Rajagopalan | CHOP | |
| Rebecca Ganetzky | CHOP | |
| Vishnu Cuddapah | CHOP | |
| Anna Raper | CHOP/UPenn | |
| Daniel J. Rader | CHOP/UPenn | |
| Giorgio Sirugo | CHOP/UPenn | |
| Vaidehi Jobanputra | Columbia | |
| Allyn McConkie-Rosell | Duke | |
| Kelly Schoch | Duke | |
| Mohamad Mikati | Duke | |
| Nicole M. Walley | Duke | |
| Rebecca C. Spillmann | Duke | |
| Vandana Shashi | Duke | |
| Alan H. Beggs | Harvard | |
| Calum A. MacRae | Harvard | |
| David A. Sweetser | Harvard | |
| Deepak A. Rao | Harvard | |
| Edwin K. Silverman | Harvard | |
| Elizabeth L. Fieg | Harvard | |
| Frances High | Harvard | |
| Gerard T. Berry | Harvard | |
| Ingrid A. Holm | Harvard | |
| J. Carl Pallais | Harvard | |
| Joan M. Stoler | Harvard | |
| Joseph Loscalzo | Harvard | |
| Lance H. Rodan | Harvard | |
| Laurel A. Cobban | Harvard | |
| Lauren C. Briere | Harvard | |
| Matthew Coggins | Harvard | |
| Melissa Walker | Harvard | |
| Richard L. Maas | Harvard | |
| Susan Korrick | Harvard | |
| Jessica Douglas | Harvard | |
| AudreyStephannie C. Maghiro | Harvard DMCC | |

| | | |
|---|---|---|
| Cecilia Esteves | Harvard DMCC | |
| Emily Glanton | Harvard DMCC | |
| Isaac S. Kohane | Harvard DMCC | |
| Kimberly LeBlanc | Harvard DMCC | |
| Rachel Mahoney | Harvard DMCC | |
| Shamil R. Sunyaev | Harvard DMCC | |
| Shilpa N. Kobren | Harvard DMCC | |
| Brett H. Graham | IU | |
| Erin Conboy | IU | |
| Francesco Vetrini | IU | |
| Kayla M. Treat | IU | |
| Khurram Liaqat | IU | |
| Lili Mantcheva | IU | |
| Stephanie M. Ware | IU | |
| Breanna Mitchell | Mayo Clinic | |
| Brendan C. Lanpher | Mayo Clinic | |
| Devin Oglesbee | Mayo Clinic | |
| Eric Klee | Mayo Clinic | |
| Filippo Pinto e Vairo | Mayo Clinic | |
| Ian R. Lanza | Mayo Clinic | |
| Kahlen Darr | Mayo Clinic | |
| Lindsay Mulvihill | Mayo Clinic | |
| Lisa Schimmenti | Mayo Clinic | |
| Queenie Tan | Mayo Clinic | |
| Surendra Dasari | Mayo Clinic | |
| Adriana Rebelo | Miami | |
| Carson A. Smith | Miami | |
| Deborah Barbouth | Miami | |
| Guney Bademci | Miami | |
| Joanna M. Gonzalez | Miami | |
| Kumarie Latchman | Miami | |
| LéShon Peart | Miami | |
| Mustafa Tekin | Miami | |
| Nicholas Borja | Miami | |
| Stephan Zuchner | Miami | |
| Stephanie Bivona | Miami | |
| Willa Thorson | Miami | |
| Herman Taylor | Morehouse DMCC | |
| Andrea Gropman | NIH UDP | |

| | | |
|---|---|---|
| Barbara N. Pusey Swerdzewski | NIH UDP | |
| Camilo Toro | NIH UDP | |
| Colleen E. Wahl | NIH UDP | |
| Donna Novacic | NIH UDP | |
| Ellen F. Macnamara | NIH UDP | |
| John J. Mulvihill | NIH UDP | |
| Maria T. Acosta | NIH UDP | |
| Precilla D'Souza | NIH UDP | precilla.d' |
| Valerie V. Maduro | NIH UDP | |
| Ben Afzali | NIH UDP, NHGRI | |
| Ben Solomon | NIH UDP, NHGRI | |
| Cynthia J. Tifft | NIH UDP, NHGRI | |
| David R. Adams | NIH UDP, NHGRI | |
| Elizabeth A. Burke | NIH UDP, NHGRI | |
| Francis Rossignol | NIH UDP, NHGRI | |
| Heidi Wood | NIH UDP, NHGRI | |
| Jiayu Fu | NIH UDP, NHGRI | |
| Joie Davis | NIH UDP, NHGRI | |
| Leoyklang Petcharet | NIH UDP, NHGRI | |
| Lynne A. Wolfe | NIH UDP, NHGRI | |
| Margaret Delgado | NIH UDP, NHGRI | |
| Marie Morimoto | NIH UDP, NHGRI | |
| Marla Sabaii | NIH UDP, NHGRI | |
| MayChristine V. Malicdan | NIH UDP, NHGRI | |
| Neil Hanchard | NIH UDP, NHGRI | |

| | | |
|---|---|---|
| Orpa Jean-Marie | NIH UDP, NHGRI | |
| Wendy Introne | NIH UDP, NHGRI | |
| William A. Gahl | NIH UDP, NHGRI | |
| Yan Huang | NIH UDP, NHGRI | |
| Aimee Allworth | PNW | |
| Andrew Stergachis | PNW | |
| Danny Miller | PNW | |
| Elizabeth Blue | PNW | |
| Elizabeth Rosenthal | PNW | |
| Elsa Balton | PNW | |
| Emily Shelkowitz | PNW | |
| Eric Allenspach | PNW | |
| Fuki M. Hisama | PNW | |
| Gail P. Jarvik | PNW | |
| Ghayda Mirzaa | PNW | |
| Ian Glass | PNW | |
| Kathleen A. Leppig | PNW | |
| Katrina Dipple | PNW | |
| Mark Wener | PNW | |
| Martha Horike-Pyne | PNW | |
| Michael Bamshad | PNW | |
| Peter Byers | PNW | |
| Sam Sheppeard | PNW | |
| Sirisak Chanprasert | PNW | |
| Virginia Sybert | PNW | |
| Wendy Raskind | PNW | |
| Nitsuh K. Dargie | PNW | |
| Beth A. Martin | Stanford | |
| Chloe M. Reuter | Stanford | |
| Devon Bonner | Stanford | |
| Elijah Kravets | Stanford | |
| Holly K. Tabor | Stanford | |
| Jacinda B. Sampson | Stanford | |
| Jason Hom | Stanford | |
| Jennefer N. Kohler | Stanford | |
| Jonathan A. Bernstein | Stanford | |
| Kevin S. Smith | Stanford | |
| Matthew T. Wheeler | Stanford | |

| | | |
|---|---|---|
| Meghan C. Halley | Stanford | |
| Page C. Goddard | Stanford | |
| Paul G. Fisher | Stanford | |
| Rachel A. Ungar | Stanford | |
| Raquel L. Alvarez | Stanford | |
| Shruti Marwaha | Stanford | |
| Terra R. Coakley | Stanford | |
| Euan A. Ashley | Stanford DMCC | |
| Ali Al-Beshri | UAB | |
| Anna Hurst | UAB | |
| Bruce Korf | UAB | |
| Kaitlin Callaway | UAB | |
| Martin Rodriguez | UAB | |
| Tammi Skelton | UAB | |
| Andrew B. Crouse | UAB DMCC | |
| Jordan Whitlock | UAB DMCC | |
| Mariko Nakano-Okuno | UAB DMCC | |
| Matthew Might | UAB DMCC | |
| William E. Byrd | UAB DMCC | |
| Changrui Xiao | UCI/CHOC | |
| Eric Vilain | UCI/CHOC | |
| Jose Abdenur | UCI/CHOC | |
| Kathyrn Singh | UCI/CHOC | |
| Rebekah Barrick | UCI/CHOC | |
| Sanaz Attaripour | UCI/CHOC | |
| Suzanne Sandmeyer | UCI/CHOC | |
| Tahseen Mozaffar | UCI/CHOC | |
| Albert R. La Spada | UCI/CHOC | |
| Elizabeth C. Chao | UCI/CHOC | |
| Maija-Rikka Steenari | UCI/CHOC | |
| Alden Huang | UCLA | |
| Brent L. Fogel | UCLA | |
| Esteban C. Dell'Angelica | UCLA | |
| George Carvalho | UCLA | |
| Julian A. Martínez-Agosto | UCLA | |
| Manish J. Butte | UCLA | |
| Martin G. Martin | UCLA | |
| Naghmeh Dorrani | UCLA | |
| Neil H. Parker | UCLA | |
| Rosario I. Corona | UCLA | |
| Stanley F. Nelson | UCLA | |

| | | |
|---|---|---|
| Yigit Karasozen | UCLA | |
| Aaron Quinlan | University of Utah | |
| Alistair Ward | University of Utah | |
| Ashley Andrews | University of Utah | |
| Corrine K. Welt | University of Utah | |
| Dave Viskochil | University of Utah | |
| Erin E. Baldwin | University of Utah | |
| John Carey | University of Utah | |
| Justin Alvey | University of Utah | |
| Laura Pace | University of Utah | |
| Lorenzo Botto | University of Utah | |
| Nicola Longo | University of Utah | |
| Paolo Moretti | University of Utah | |
| Rebecca Overbury | University of Utah | |
| Russell Butterfield | University of Utah | |
| Steven Boyden | University of Utah | |
| Thomas J. Nicholas | University of Utah | |
| Matt Velinder | University of Utah | |
| Gabor Marth | University of Utah DMCC | |
| Pinar Bayrak-Toydemir | University of Utah/ARUP | |
| Rong Mao | University of Utah/ARUP | |
| Monte Westerfield | UO MOSC | |
| Brian Corner | Vanderbilt | |

| | | |
|---|---|---|
| John A. Phillips III | Vanderbilt | |
| Kimberly Ezell | Vanderbilt | |
| Lynette Rives | Vanderbilt | |
| Rizwan Hamid | Vanderbilt | |
| Serena Neumann | Vanderbilt | |
| Ashley McMinn | Vanderbilt | |
| Joy D. Cogan | Vanderbilt | |
| Thomas Cassini | Vanderbilt | |
| Alex Paul | WUSTL Clinical | |
| Dana Kiley | WUSTL Clinical | |
| Daniel Wegner | WUSTL Clinical | |
| Erin McRoy | WUSTL Clinical | |
| Jennifer Wambach | WUSTL Clinical | |
| Kathy Sisco | WUSTL Clinical | |
| Patricia Dickson | WUSTL Clinical | |
| F. Sessions Cole | WUSTL DMCC | |
| Dustin Baldridge | WUSTL MOSC | |
| Jimann Shin | WUSTL MOSC | |
| Lilianna Solnica-Krezel | WUSTL MOSC | |
| Stephen Pak | WUSTL MOSC | |
| Timothy Schedl | WUSTL MOSC | |
| Hector Rodrigo Mendez | Stanford | |
| Brianna Tucker | Stanford | |
| Beatriz Anguiano | Stanford | |
| Mia Levanto | Stanford | |
| Suha Bachir | Stanford | |
| Laurens Wiel | Stanford | |
| Stephen B Montgomery | Stanford | |
| Tanner D Jensen | Stanford | |
| John E. Gorzynski | Stanford | |
| Sara Emami | Stanford | |
| Laura Keehan | Stanford | |
| Jennifer Schymick | Stanford | |
| Taylor Maurer | Stanford | |
| Alexander Miller | Stanford | |
| Andres Vargas | UCLA | |
| Amanda M. Shrewsbury | UCLA | |
| Bianca E. Russell | UCLA | |
| Layal F. Abi Farraj | UCLA | |
| Elizabeth A Worthey | UAB | |
| Tarun KK Mamidi | UAB | |

| | | |
|---|---|---|
| Brandon M Wilk | UAB | |
| Rachel Li | Sanford | |
| Jennifer Morgan | Sanford | |
| Chun-Hung Chan | Sanford | |
| Paul Berger | Sanford | |
| Mohamad Saifeddine | Sanford | |
| Isum Ward | Sanford | |
| Jason Schend | Sanford | |
| Megan Bell | Sanford | |
| Dr. Francisco Bustos velasq | Sanford | |
| Taylor Beagle | Sanford | |
| Miranda Leitheiser | Sanford | |
| Runjun Kumar | PNW | |
| Donald Basel | MCW-CW | |
| Michael Muriello | MCW-CW | |
| Brett Bordini | MCW-CW | |
| Michael Zimmermann | MCW-CW | |
| Abdul Elkadri | MCW-CW | |
| James Verbsky | MCW-CW | |
| Julie McCarrier | MCW-CW | |

Members of Baylor College of Medicine Center for Precision Medicine Models

| Names | Affiliation(s) |
|---|---|
| Aleksander Milosavljevic | 1) Department of Molecular and Human Genetics, BCM |
| Matthew Roth | 1) Department of Molecular and Human Genetics, BCM |
| Ramin Zahedi Darshoori | 1) Department of Molecular and Human Genetics, BCM |
| Uma Ramamurthy | 1) Office of Research Information Technology |
| Vivek Ramanathan | 1) Office of Research Information Technology |
| Zhandong Liu | 1) Department of Pediatrics, BCM; 2) Jan and Dan Duncan Neurological Research Institute, Texas Children's Hospital |
| Seon-Young Kim | 1) Department of Pediatrics, BCM; 2) Jan and Dan Duncan Neurological Research Institute, Texas Children's Hospital |
| Athanasios Papastathopoulos-Katsaros | 1) Department of Pediatrics, BCM; 2) Jan and Dan Duncan Neurological Research Institute, Texas Children's Hospital |
| Lindsay C Burrage | 1) Department of Molecular and Human Genetics, BCM |
| Jill A Rosenfeld | 1) Department of Molecular and Human Genetics, BCM |
| Sandesh CS Nagamani | 1) Department of Molecular and Human Genetics, BCM |
| Jennifer E Posey | 1) Department of Molecular and Human Genetics, BCM |
| Jason D Heaney | 1) Department of Molecular and Human Genetics, BCM; 2) Dan L Duncan Comprehensive Cancer Center, BCM |
| Denise G Lanza | 1) Department of Molecular and Human Genetics, BCM |
| Shinya Yamamoto | 1) Department of Molecular and Human Genetics, BCM; 2) Department of Neuroscience, BCM, 3) Jan and Dan Duncan Neurological Research Institute, Texas Children's Hospital |
| Michael F Wangler | 1) Department of Molecular and Human Genetics, BCM; 2) Jan and Dan Duncan Neurological Research Institute, Texas Children's Hospital |
| Hugo J Bellen | 1) Department of Molecular and Human Genetics, BCM; 2) Department of Neuroscience, BCM, 3) Jan and Dan Duncan Neurological Research Institute, Texas Children's Hospital |
| Oguz Kanca | 1) Department of Molecular and Human Genetics, BCM; 2) Jan and Dan Duncan Neurological Research Institute, Texas Children's Hospital |
| Jeffrey Rogers | 1) Department of Molecular and Human Genetics, BCM; 2) Human Genome Sequencing Center, BCM |
| Muthuswamy Raveendran | 1) Department of Molecular and Human Genetics, BCM; 2) Human Genome Sequencing Center, BCM |
| Sharayu Jangam | 1) Department of Molecular and Human Genetics, BCM; 2) Jan and Dan Duncan Neurological Research Institute, Texas Children's Hospital |
